# Large deletions perturb peripheral transcriptomic and metabolomic profiles in Phelan-McDermid syndrome

**DOI:** 10.1101/2022.07.06.22277334

**Authors:** Michael S. Breen, Xuanjia Fan, Tess Levy, Rebecca Pollak, Brett Collins, Aya Osman, Anna S. Tocheva, Mustafa Sahin, Elizabeth Berry-Kravis, Latha Soorya, Audrey Thurm, Craig M. Powell, Jonathan A. Bernstein, Alexander Kolevzon, Joseph D. Buxbaum, the Developmental Synaptopathies Consortium

## Abstract

Phelan-McDermid syndrome (PMS) is a rare neurodevelopmental disorder caused at least in part by haploinsufficiency of the SHANK3 gene, due to sequence variants in SHANK3 or subtelomeric 22q13.3 deletions. Phenotypic differences have been reported between PMS participants carrying small ‘Class I’ mutations and large ‘Class II’ mutations, however the molecular perturbations underlying these divergent phenotypes remain obscure. Using peripheral blood transcriptome and serum metabolome profiling, we examined the molecular perturbations in the peripheral circulation associated with a full spectrum of PMS genotypes spanning Class I (*n*=37) and Class II mutations (*n*=39). Transcriptomic data revealed 52 genes with blood expression profiles that tightly scale with 22q.13.3 deletion size. Further, we uncover 208 under-expressed genes in PMS participants with Class II mutations, which were unchanged in Class I mutations. These genes were not linked to 22q13.3 and were strongly enriched for glycosphingolipid metabolism, NCAM1 interactions and cytotoxic natural killer (NK) immune cell signatures. *In silico* predictions estimated a reduction in CD56+ CD16-NK cell proportions in Class II mutations, which was validated by mass cytometry time of flight. Global metabolomics profiling identified 24 metabolites that were significantly altered with PMS participants with Class II mutations, and confirmed a general reduction in sphingolipid metabolism. Collectively, these results provide new evidence linking PMS participants carrying Class II mutations with decreased expression of cytotoxic cell signatures, reduced relative proportions of NK cells, and lower sphingolipid metabolism. These findings highlight alternative avenues for therapeutic development and offer new mechanistic insights supporting genotype-to-phenotype associations in PMS.

## BACKGROUND

Phelan-McDermid syndrome (PMS) is one of the most penetrant and common single-locus causes of autism spectrum disorder (ASD) and accounts for ca. 1% of ASD diagnoses^1-3^. PMS is caused by heterozygous 22q13.3 deletions or *SHANK3* sequence variants leading to haploinsufficiency of the *SHANK3* gene^2-6^. Participants with PMS present with a constellation of clinical and neurobehavioral phenotypes, including neonatal hypotonia, global developmental delay, intellectual disability (ID), severely delayed or absent speech, and/or frequent ASD^6-8^. Additional features can also include seizures, motor skill deficits, and structural brain abnormalities^8^. Heterogeneity in the clinical presentation of PMS is not fully explained by sequence variants or deletions limited to the *SHANK3* locus, emphasizing the importance of understanding the broader genetic landscape of PMS.

The majority of reported cases of PMS are caused by large 22q13.3 deletions, which encompass additional genes and can extend up to 9.2 Mb^6-9^. Given the variable nature of the deletions, it is useful to classify PMS genotypes as either Class I mutations (including SHANK3 sequence variants or deletions in *SHANK3* only or *SHANK3* with *ARSA* and/or *ACR* and *RABL2B*), or Class II mutations (all other deletions)^8^. The largest genotype-phenotype association analysis indicates that PMS participants with Class II mutations display increased rates of early developmental delays, intellectual disability, minimally verbal status, and various medical features^8^. Notably, individuals with Class I mutations attained more advanced developmental milestones, which were reached at a younger age compared to those with Class II mutations, and were more likely to exhibit higher language and communication skills^8^. These results are largely consistent with smaller independent reports^6,7,10^, and together emphasize that the frequency and severity of PMS phenotypes is likely caused by haploinsufficiency of multiple additional candidate genes. A next practical step would be to identify consistent molecular changes resulting from these specific genetic alterations in individuals with PMS.

SHANK3 is a scaffolding protein of the postsynaptic density of glutamatergic synapses^11-13^; additional disrupted genes within larger Class II mutations have been implicated in processes related to stress and inflammation, mitochondrial function, neuronal differentiation, and cellular metabolism^14-17^. Molecular profiling of tissues derived from PMS participants, albeit scarce, confirm these functional categories and support the notion of unique molecular programs underlying distinct clinical subtypes. For example, increased severity of PMS phenotypes and larger 22q13.3 deletions have been associated with alterations in mitochondrial complex I and IV activity^15^, changes in peripheral blood epi-signatures enriched for neuronal development and intracellular signaling^16^, and metabolomic changes implicated in metabolic stress and response to cytokines regulating inflammation^16-17^, all poised to influence neurodevelopment. Notably, even transcriptomics of peripheral blood and postmortem brain tissue from participants with idiopathic ASD implicate changes related to inflammation, cellular proliferation/metabolism and immune dysfunction^18-20^, supporting the notion that ongoing dysregulation of the immune system echoes alterations in the central nervous system (CNS). Despite these advances, studies examining the molecular changes underlying specific genetic alterations in PMS commonly employ modest sample sizes, implement variable assessment methods for measuring clinical phenotypes, and apply differing thresholds for defining large and small deletions, making it challenging to elucidate the full spectrum of genes and pathways associated with genes disrupted on 22q13.3.

Given the success of blood transcriptome profiling to identify novel mechanisms and high-confidence targets for several rare CNS disorders^21-24^, including idiopathic ASD^20^ more broadly, we hypothesized that unbiased peripheral blood transcriptomic and metabolomic profiling across a large spectrum of genotypes would shed light on the molecular changes underlying specific genetic alterations in PMS.

The objective of the current study was to examine the molecular perturbations in the peripheral circulation associated with a full spectrum of PMS genotypes. A total of 76 PMS probands were included in the current study. Peripheral blood transcriptomic data were generated across 68 PMS participants, including Class I mutations (*n*=33) and Class II mutations (*n*=35), as well as an age and sex matched control group (*n*=24). Additionally, global metabolomic data were generated across a partially overlapping subset of 25 PMS participants, comprised of Class I mutations (*n*=11), Class II mutations (*n*=14), and an age and sex matched control group (*n*=29). Using a combination of genotypic, transcriptomic and metabolomics data, we sought to: i) elucidate key genes, pathways and cell types altered in PMS participants with Class I and Class II mutations; ii) explore molecular relationships between gene expression patterns and clinical features of PMS; and iii) identify core sets of differentially abundant metabolites in PMS participants with Class I and Class II mutations. We identify a molecular footprint of Class II mutations, which informs pathobiological mechanisms in PMS and suggest approaches for interventions.

## METHODS

### Ascertainment of PMS participants and collection of clinical phenotypes

Informed consent was obtained from participants’ caregivers for study participation, as previously described^8^. The cohort included 76 PMS participants (38 female, 38 male) between the ages of one and 42 years (8.9 ± 6.5). Forty-six participants were enrolled in studies at the Seaver Autism Center for Research and Treatment at the Icahn School of Medicine at Mount Sinai. An additional 30 participants were enrolled by partner sites through the Rare Disease Clinical Research Network Developmental Synaptopathies Consortium (DSC), as part of a PMS phenotyping and natural history study. For each participant, a comprehensive battery of standardized assessments, semi-structured interviews, and caregiver report questionnaires was used to examine medical comorbidities, intellectual and adaptive functioning, expressive and receptive language, ASD symptomatology, and behavioral comorbidities, as previously described^8^. Studies were approved by the Institutional Review Board (IRB) for the protection of human subjects at Mount Sinai (Study IDs: 98–0436, 10-0527, 12-1718) and Boston Children’s Hospital (Study ID: P00013300), which serves as the central IRB for the DSC.

### Peripheral blood RNA isolation, library preparation and quantification of gene expression

Peripheral blood was collected in PAXgene Blood RNA tubes (Qiagen, Valencia, CA, USA) for 68 PMS participants. Peripheral blood was also collected from 24 unaffected control subjects (12 female, 12 male) between the ages of one and 24 years (9.5 ± 4.9), 21 of which were unaffected familial siblings. Total RNA was extracted and purified in accordance with the PAXgene Blood RNA Kit instructions (Qiagen, Valencia, CA, USA). Globin mRNA was depleted from samples using the GLOBINclear Human Kit (Life Technologies, Carlsbad, CA, USA). The quantity of purified RNA was measured on a Nanodrop 2000 Spectrophotomerter (Thermo Scientific; 61.4 ± 24.1 ng µl^-1^) and RNA integrity numbers (RIN) measured with the Agilent 2100 Bioanalyzer (Agilent, Santa Clara, CA, USA; 8.0 ± 0.3). The Illumina TruSeq Total RNA kit (Illumina, San Diego, CA, USA) was used for library preparation accordingly to manufacturer instructions without any modifications. Indexed RNA libraries were pooled and sequenced using long paired-end chemistry (2×150 bp) at an average read depth of ∼11M reads per sample using the Illumina HiSeq2500. All high-quality trimmed reads were mapped to UCSC *Homo sapiens* reference genome (build hg37) using default STAR v2.5.3 parameters^24^. Samtools was used to convert bamfiles to samfiles and featureCounts^25^ was used to quantify gene expression levels for each individual sample using default paired-end parameters.

### RNA-seq data quality control

Raw count data measured 56,632 genes across 92 participants. Unspecific filtering removed lowly expressed genes that did not meet the requirement of a minimum of 1 count per million (cpm) in at least 15 subjects (∼16% of subjects). A total of 16,285 genes were retained and defined as stably expressed in peripheral blood. These genes were subjected to limma VOOM normalization^26^ and inspected for outlying samples using unsupervised hierarchical clustering of subjects (based on Pearson coefficient and average distance metric) and principal component analysis to identify potential outliers outside two standard deviations from these averages. No such outliers were identified in the current dataset.

### Gene-based annotations for loss of function intolerance

We collected probability of loss of function (LoF) intolerance (pLI) scores from the gnomAD project (https://gnomad.broadinstitute.org/). pLI scores indicate whether a gene is intolerant for either heterozygous or homozygous LoF variants, and was used to classify disrupted genes on 22q13.3 as either definitely LoF intolerant (pLI ≥ 0.9), possible LoF intolerant (0.5 ≥ pLI < 0.9) or definitely LoF tolerant (pLI ≤ 0.1).

### Differential gene expression and association testing

A moderated *t*-test implemented through the *limma* package^26^ was used to assess differential gene expression between unaffected controls and three different groupings of PMS participants: 1) all PMS participants; 2) Class I mutations only; and 3) Class II mutations only. We also 4) tested for differences between participants with Class II mutations and Class I mutations. These analyses tested PMS genotypes as the primary main outcome. Subsequently, we performed a secondary exploratory analysis examining relationships between gene expression and 19 clinical phenotypes within PMS participants only. Each clinical phenotype was tested separately and any participant with missing data would be dropped from the analysis, respectively. All analysis described here covaried for the possible influence of sex and age on gene expression differences. Significance threshold was set to a Benjamini-Hochberg (BH) multiple test corrected *P*-value <0.05 to control the false discovery rate (FDR), unless specified otherwise.

### Functional annotation of differentially expressed genes

Correlation adjusted mean rank (CAMERA) gene set enrichment was performed using the resulting sets of summary statistics^26,27^. CAMERA performs a competitive gene set rank test to assess whether the genes in a given gene-set are highly ranked in terms of differential expression relative to genes that are not in the gene-set. For example, the test ranks gene expression differences in PMS participants with Class II mutations relative to unaffected controls to test whether gene-sets are over-represented towards the extreme ends of this ranked list. After adjusting the variance of the resulting gene-set test statistic by a variance inflation factor that depends on the gene-wise correlation (which we set to default parameters, 0.01) and the size of the set, a *p*-value is returned and adjusted for multiple testing. We used this function to test two aims: First, we examined each resulting set of PMS-associated changes in gene expression for enrichment of biological processes and pathways using a well-curated collection of REACTOME pathways and Gene Ontology Molecular Factors (GO:MF). We specifically focused on functional annotation of differentially expressed genes *i)* across all PMS participants, ii) participants with Class I mutations only; iii) participants with Class II mutations only; and iv) changes between Class I and Class II mutations.

### Cell type-specific gene set enrichment analysis using scRNA-seq data

Three single-cell RNA-sequencing (scRNA-seq) experiments were downloaded and incorporated in the current study: the first dataset comprised of 10,975 PBMCs (v2 Chemistry) and the second dataset comprised of 33,227 PBMCs (v2 Chemistry), both were downloaded from the list of publically available 10X Genomic Inc. datasets; the third data set was comprised of 67,272 PBMCs and was obtained from Zheng et al., 2017^28^. For each dataset, we used pre-computed filtered, normalized, and scaled data together with pre-existing cell type classifications as originally described and deposited for each data set. Thus, no additional data processing was performed as each experiment was pre-processed and quality controlled. Next, cell type marker genes were curated across all three experiments using the FindAllMarkers function in the Seruat R package^29^ with the following specifications: min.pct = 0.25, logfc threshold = 0.01, FDR *p*-value < 0.05. These resulting lists of cell type markers were compiled into cell type-specific gene sets and used as input to perform CAMERA gene-set enrichment analysis (as described above) to determine if a rank ordered list of PMS-related differentially expressed genes contained an over-representation of cell type-specific genes towards either extreme ends. A separate independent cell-type enrichment analysis was performed using the three datasets. Rather than testing for the distribution of cell type-specific marker genes along a ranked list of PMS-related genes, we directly queried the expression of a given list of PMS-related genes within and across all individual single cells using singular value decomposition. Thus, the expression of each PMS-related gene set was aggregated into one singular eigengene value, which was plotted within and across all single cells as a global representative of gene expression for a given gene set of interest.

### *In silico* cytometry estimates the proportions of peripheral blood immune cells

The frequencies of circulating blood immune cells were estimated for each individual in each study using CIBERSORTx cell type de-convolution (https://cibersortx.stanford.edu/)^30^. CIBERSORTx relies on known cell subset specific marker genes and applies linear support vector regression, a machine learning approach highly robust compared to other methods with respect to noise, unknown mixture content and closely related cell types. As input, we used the LM22 signature matrix to distinguish nine main leukocytes subtypes: B cells (CD19+), T cells (CD3+), natural killer (NK) cells (CD56+), monocytes (CD14+), dendritic cells, mast cells, macrophages, eosinophils and neutrophils. The means of the resulting estimates were compared between PMS participants and unaffected controls and tested for significance using a Student’s *t*-test.

### Cytometry by time of flight (CyTOF): data acquisition, pre-processing, and analysis

High dimensional immuno-phenotyping by CyTOF was performed on frozen stabilized peripheral blood mononuclear cells (PBMCs) from five PMS participants with Class II mutations and four age matched pediatric control participants. Thawed PBMCs were delivered to the Human Immune Monitoring Core at the Icahn School of Medicine at Mount Sinai in fresh RPMI media. Samples were washed in Cell Staining Buffer (CBS; Fluidigm, San Francisco, CA, USA) and re-suspended in fresh CSB. Fc Receptor blocking (Biolegend, San Diego, CA, USA), Rh103 viability staining (Fluidigm), and live-cell barcoding were all performed simultaneously at room temperature (citations). After a 30-minute incubation at room temperature, samples were washed twice in CSB, pooled, and stained with surface markers for 30 minutes at room temperature. Two CSB washes were performed. Samples were then fixed with 2.4% PFA and subsequently labeled with Iridium and Osmium for 30 minutes at room temperature. Samples were washed twice in CSB and stored in CSB until acquisition.

Prior to data acquisition, samples were washed in Cell Acquisition Solution (Fluidigm) and resuspended at a concentration of 1 million cells per ml in Cell Acquisition Solution containing a 1:20 dilution of EQ Normalization beads (Fluidigm). The samples were then acquired on a Helios Mass Cytometer equipped with a wide-bore sample injector at an event rate of <400 events per second. After acquisition, repeat acquisitions of the same sample were concatenated and normalized using the Fluidigm software and uploaded to Cytobank for data analysis.

Cells were first identified based on Ir-193 DNA intensity and CD45 expression; Ce140+ normalization beads, CD45-low/Ir-193-low debris and cross-sample and Gaussian ion-cloud multiplets were excluded from downstream analysis. After this data cleanup, manual gating was utilized to debarcode the multiplexed live-cell barcoded sample. The FCS files were split by debarcoded population to complete debarcoding and data clean-up. The cell counts and frequencies of the annotated cell subsets, excluding debris and known cell–cell multiplets, were exported for downstream statistical analyses. To identify changes in cellular populations we performed differential abundance analysis using a moderated *t*-test implemented through limma^26^. The annotated cell frequencies were used as input into a model fit using Class II mutations as the outcome variable.

### Global plasma metabolomics profiling and data pre-processing

Plasma was isolated from 54 participants (*n*=29 unaffected controls; *n*=11 Class I mutations; *n*=14 Class II mutations) by centrifugation of blood samples in EDTA tubes for 30 min at 1,500*g*. Notably, 17 PMS participants and 12 unaffected controls had paired peripheral blood transcriptomic data. Separated plasma aliquots of 0.5 ml were stored immediately at -80 °C until transport in dry ice for global metabolomic profiling using the analytical DiscoveryHD4 platform by Metabolon, as previously described^31,32^. Raw data were extracted and signature chromatographic peaks and relative ion concentrations for metabolites detected were identified for each sample. Spectrometry data were analyzed using the Quantify Individual Components in a Sample method^32^. Metabolite identification was performed by matching each metabolite aggregate to an annotated reference chemical library containing >4,000 metabolites with well-defined chemical profiles. Peaks were quantified using the area under the curve. Metabolite data were then normalized in terms of raw peak area counts and re-scaled to set the median equal to one. Subsequently, any missing values, which constituted ∼8% of the entire data frame, were imputed with the minimum. Finally, we removed metabolites with low standard deviation (< 0.01) across the entire cohort, yielding 1,045 metabolites.

### Metabolomics statistical analyses

A moderated *t*-test from *limma*^26^ was used to assess differential abundance of metabolites between unaffected controls and three different groupings of PMS participants: 1) all PMS participants; 2) Class I mutations only; and 3) Class II mutations only. We also 4) tested for differences between participants with Class II mutations and Class I mutations. These analyses adjusted for the possible influence of sex and age on metabolite profiles and a significance threshold was set to a BH multiple test corrected *P*-value < 0.1 to control the FDR. Differentially abundant metabolites were subjected to pathway annotation using MetaboAnalyst5.0 (https://www.metaboanalyst.ca)^33^. We applied a joint pathway analysis to integrate our transcriptomic and metabolomic data and interpret them at a pathway level. To do so, the mass of each metabolite detected was queried against the Human Metabolome Database (HMDB)^34^. Once identified, a list of differentially expressed genes and differentially abundant metabolites identified by the HMDB was imported into MetaboAnalyst5.0 along with their direction of effect (log_2_ fold-changes). These results mapped to well-curated molecular pathways for over-representation analysis using hypergeometric tests, and *P*-values were adjusted using Holm–Bonferroni correction.

## RESULTS

### Clinical features of PMS participants with Class I and Class II mutations

A total of 76 PMS probands were included in the current study. (**Table 1**). Across the full cohort, 17 participants had sequence variants in *SHANK3*, including 13 frameshift, 2 nonsense, one splice site and one *de novo* missense variant (**Figure S1**). Participants were parsed into two groups: 1) Class I mutations: sequence variants or small deletions including only *SHANK3 or SHANK3* in combination with *ARSA* and/or *ACR* and *RABL2B;* and 2) Class II mutations: all larger deletions that did not qualify as Class I mutations. Participants with Class II mutations exhibit significantly lower full-scale, verbal and nonverbal IQ/DQ relative to Class I mutations (*p*=0.045, *p*=0.023, *p*=0.019, respectively), consistent with existing evidence for genotype-phenotype associations in PMS^8^. Notably, Class II mutations also display significantly reduced motor skills on the Vineland-2 Adaptive Behavior Scale (VABS Motor), but were also younger when compared to participants with Class I mutations (*p*=0.006, *p*=0.02, respectively). A subset of PMS participants in the current study underwent peripheral blood transcriptome profiling (*n*=68) and/or serum metabolomic profiling (*n*=25) (**Table S1**), which were the focus of the subsequent analyses.

**Table 1.** Clinical features of Class II and Class I mutations in the current study.

|  | *Entire cohort of 76 PMS probands* |  |  |  | *Transcriptome subset (68 probands)* |  | *Metabolome subset (25 probands)* |  |
| --- | --- | --- | --- | --- | --- | --- | --- | --- |
|  | Class II (n=39) | Class I (n=37) | P-value | Effect size | P-value | Effect size | P-value | Effect size |
| Sex, M/F | 16/23 | 22/15 | 0.169 | -0.180 | 1.000 | 0.010 | 0.877 | -0.120 |
| Caucasian/Other | 33/6 (84%) | 33/4 (89%) | 0.802 | -0.070 | 1.000 | -0.010 | 0.565 | 0.250 |
| Hispanic/Other | 4/35 (11%) | 4/33 (12%) | 1.000 | -0.010 | 0.215 | -0.180 | 0.363 | -0.250 |
| Age (months) | 85.821 ± 47.781 | 129.432 ± 97.734 | 0.026 | -0.579 | 0.039 | -0.564 | 0.317 | -0.180 |
| Verbal IQ/DQ | 22.354 ± 17.494 | 33.617 ± 24.181 | 0.045 | -0.546 | 0.034 | -0.645 | 0.305 | -0.490 |
| Nonverbal IQ/DQ | 28.688 ± 17.237 | 39.973 ± 21.916 | 0.023 | -0.583 | 0.017 | -0.699 | 0.222 | -0.383 |
| Full scale IQ/DQ | 24.739 ± 16.003 | 34.218 ± 19.943 | 0.019 | -0.535 | 0.016 | -0.623 | 0.272 | -0.437 |
| ADOS Total | 15.912 ± 6.122 | 15.821 ± 8.094 | 0.854 | 0.013 | 0.891 | 0.085 | 0.717 | 0.745 |
| ADOS SA | 12.941 ± 5.116 | 11.882 ± 6.741 | 0.676 | 0.180 | 0.464 | 0.276 | 0.802 | 0.880 |
| ADOS RRB | 3 ± 1.969 | 3.939 ± 2.318 | 0.066 | -0.444 | 0.063 | -0.463 | 0.431 | -0.475 |
| ADI Communication | 12.697 ± 4.355 | 12.065 ± 4.761 | 0.425 | 0.141 | 0.425 | 0.070 | 0.410 | 1.401 |
| ADI Social | 19.529 ± 6.872 | 17.031 ± 8.656 | 0.313 | 0.326 | 0.359 | 0.308 | 0.094 | 1.880 |
| ADI RRB | 4.771 ± 2.426 | 4.594 ± 3.12 | 0.743 | 0.065 | 0.822 | 0.050 | 0.451 | 1.241 |
| VABS Communication | 50.564 ± 15.441 | 52.722 ± 19.823 | 0.531 | -0.124 | 0.268 | -0.333 | 0.795 | -0.949 |
| VABS DLS | 51.821 ± 13.483 | 53.278 ± 16.77 | 0.702 | -0.098 | 0.505 | -0.154 | 0.897 | -0.725 |
| VABS Social | 56.513 ± 12.941 | 60.75 ± 17.977 | 0.313 | -0.276 | 0.230 | -0.361 | 0.938 | -0.756 |
| VABS Motor | 55.324 ± 11.954 | 63.188 ± 10.187 | 0.006 | -0.714 | 0.004 | -0.774 | 0.051 | -0.117 |
| VABS Comp | 51.308 ± 12.404 | 55.306 ± 16.603 | 0.190 | -0.278 | 0.171 | -0.327 | 0.203 | -0.503 |
| ABC Irritability | 10.114 ± 9.536 | 8.286 ± 11.184 | 0.158 | 0.179 | 0.117 | 0.312 | 0.560 | 0.707 |
| ABC Social Withdrawal | 10.943 ± 9.142 | 9.029 ± 7.86 | 0.485 | 0.228 | 0.328 | 0.270 | 0.950 | 0.992 |
| ABC Stereotypic Behavior | 4.857 ± 4.846 | 5.371 ± 6.193 | 0.887 | -0.094 | 0.714 | 0.096 | 0.269 | 0.239 |
| ABC Hyperactivity | 19.265 ± 12.425 | 19 ± 13.083 | 0.876 | 0.021 | 0.979 | -0.027 | 0.829 | -0.741 |
| ABC Inappropriate Speech | 1.457 ± 2.091 | 3.371 ± 5.719 | 0.113 | -0.451 | 0.259 | -0.343 | 0.055 | -0.071 |
| Recurrent infections, Y/N | 13/24 (54%) | 11/22 (50%) | 1.000 | 0.012 | 1.000 | 0.010 | 1.000 | 0.010 |

Continuous measures were analyzed using a Mann Whitney-U test and effect sizes computed using Cohen's D. Discrete measures were analyzed using a Chi-Squared Test and effect sizes computed using the phi coefficient. Abbreviations: Aberrant Behavior Checklist (ABC); Autism Diagnostic Interview (ADI); Autism Diagnostic Observation Schedule (ADOS); Daily Living Skills (DLS); Developmental Quotient (DQ); Intellectual Quotient (IQ); Restrictive and Repetitive Behavior (RRB); Social Affect (SA); Vineland Adaptive Behavior Scales (VABS). Recurrent infections were clinician graded and defined as more than two pneumonia or sinus infections per year.

### Class II mutations, but not Class I mutations, alter transcriptional profiles in peripheral blood

Peripheral blood transcriptomic data were generated across 68 PMS participants, including Class I sequence variants and mutations (*n*=33) and Class II mutations (*n*=35), as well as an age and sex matched control group (*n*=24), which largely consisted of unaffected siblings (∼91%). Given the breadth of genes affected by large Class II mutations on the terminal end of the long arm of chromosome 22 (22q13.3) (**Figure S2A**), we queried which of these genes are stably expressed in peripheral blood. Of 128 genes affected by large Class II mutations, 52 genes were stably expressed and detected in peripheral blood (∼40%), including genes *ARSA, RABL2B*, and *BRD1*, which were affected by the majority of Class II mutations (**Figure 1A**). Unsupervised hierarchical clustering applied to these 52 blood-expressed genes on 22q13.3 accurately distinguished 85% of Class II mutations (30/35) from all other participants, on the basis of reduced expression levels of these genes. (**Figure 1B**). Participants with Class I mutations and unaffected controls clustered together and displayed higher expression levels on average for this subset of 22q13.3 genes. To determine which of the disrupted genes on 22q13.3 are intolerant to heterozygous and homozygous loss of function (LoF), we computed probability of LoF intolerance (pLI) scores. Using this metric, we classified eight of the 52 blood expressed genes as ‘possibly LoF intolerant’ (0.5 ≥ pLI < 0.9) and eight additional genes as ‘definitely LoF intolerant’ (pLI ≥ 0.9), including genes *TRABD, PIM3, TBC1D22A, ZBED4, PLXNB2, BRD1, GRAMD4* and *CELSR1* (**Figure S2B**). Notably, all 52 genes are broadly expressed across 30 distinct human tissues from the GTEx project (**Figure S2C**), suggesting their disruption may affect diverse biological systems across a range of tissues.

**Figure 1.**
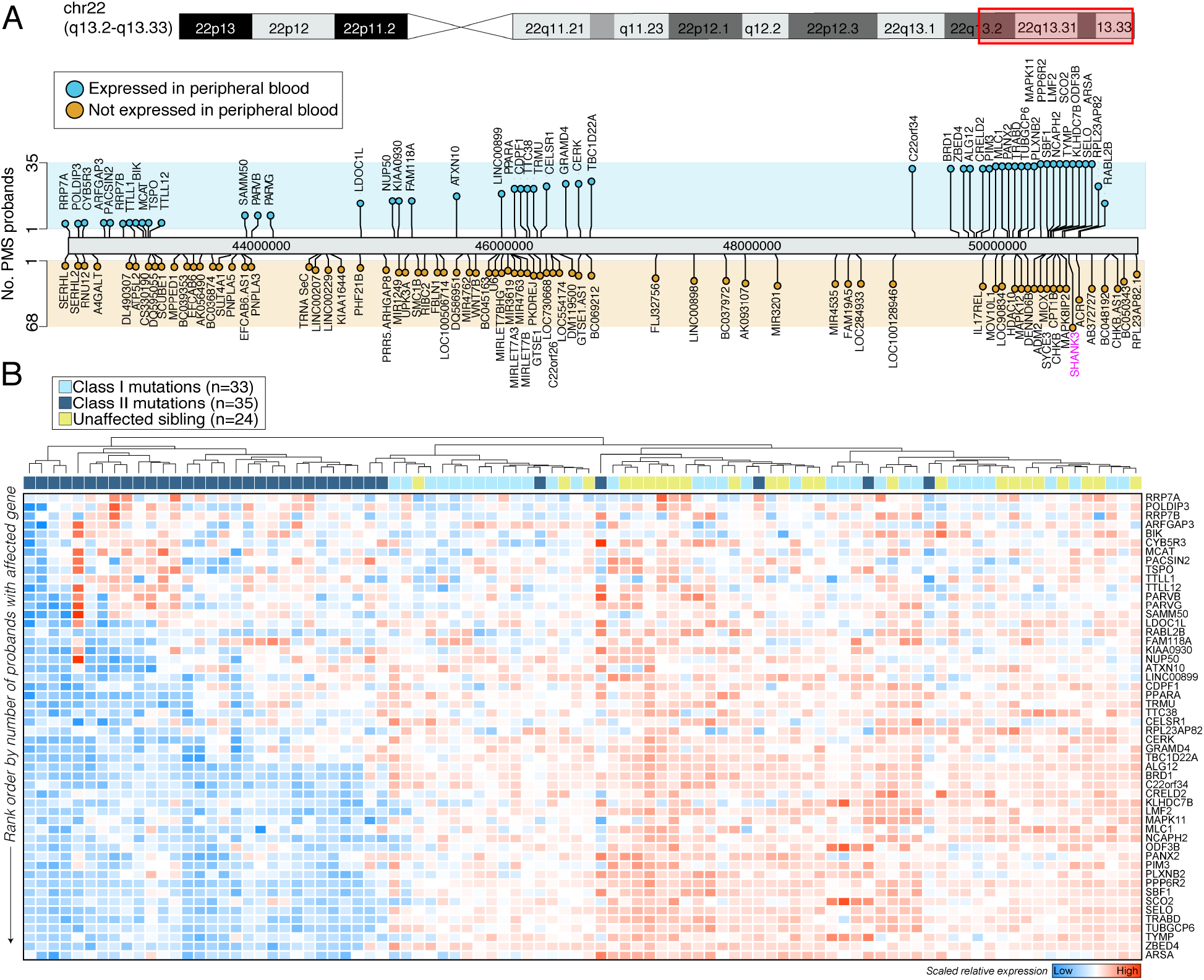
The landscape of Class I and Class II mutations in PMS. (**A**) Lollipop plot of genes affected by Class I mutations and Class II mutations across the terminal end of the long arm of chromosome 22 (22q13.3) in the 68 PMS probands included in the study. Genes are displayed as either expressed (blue; n=52 genes) in peripheral blood or not (orange; n=76 genes) and ranked by the number of probands harboring the affected gene (y-axis). *SHANK3* is highlighted in pink. (**B**) Unsupervised hierarchal clustering and heatmap (blue=low; red=high) depiction of the 52 genes on 22q13.3 that are expressed in peripheral blood affected by Class I and Class II mutations. Clustering distinguishes probands with Class II mutations from those with Class I mutations and unaffected controls. Genes were rank ordered by the number of PMS participants with the affected gene (y-axis; rare to more frequent).

Transcriptome-wide differences in gene expression were modelled for i) all PMS participants, ii) Class I mutations, and iii) Class II mutations, each relative to unaffected controls. We also modelled for iv) differences between Class II mutations and Class I mutations. Overall, the largest effect was observed between participants with Class II mutations and unaffected controls, uncovering 208 under-expressed genes and 42 over-expressed genes associated with Class II mutations (FDR < 5%) (**Figure 2A, Table S2**). Similarly, differences between Class II and Class I mutations revealed 89 under-expressed genes and 2 over-expressed associated with Class II mutations, ∼84% of these genes were also differentially expressed in Class II mutations compared to unaffected controls (**Figure S3**). There were no significant changes observed between Class I mutations compared to unaffected controls. Likewise, few genes were significantly differentially expressed when comparing all PMS participants (Class I and Class II mutations) to unaffected controls (*n*=23 genes). These results indicate that gene expression profiles are similar between participants with Class I mutations and unaffected controls, whereas participants with Class II mutations are associated with unique peripheral blood transcriptional signatures.

**Figure 2.**
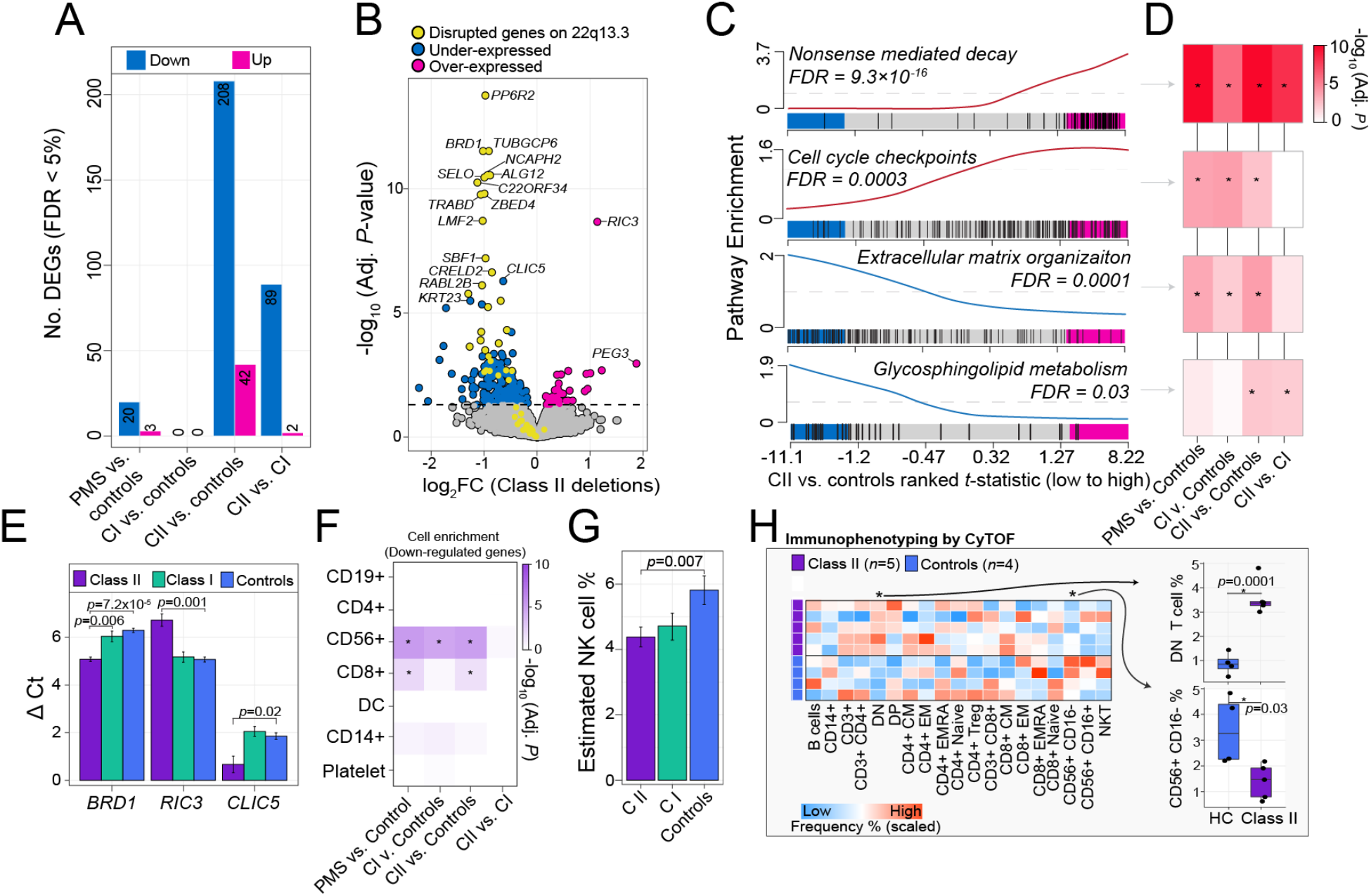
Altered peripheral blood gene expression profiles in Class II mutations. (**A**) The total number of differentially expressed genes (DEGs; y-axis) for each comparison (x-axis). Each analysis adjusted for sex and age as covariates. (**B**) Volcano plot of Class II DEGs relative to unaffected controls depicting log_2_ fold-change (log_2_FC; x-axis) and -log_10_ FDR adjusted *p*-value (y-axis). The horizontal line indicates FDR < 5%. Genes in yellow are the 52 genes expressed in blood that are affected by large Class II mutations in PMS. Genes in blue are all other down-regulated genes and those in pink are all other up-regulated genes. (**C**) Four representative pathway enrichment scores (y-axis) of Class II DEGs according to ranked t-statistics, high (pink) to low (blue) (x-axis). All enrichment results can be found in Table S2. (**D**) The resulting FDR adjusted *p*-value enrichment for all differential comparisons reveals shared and unique gene set enrichment among Class II and Class I mutations. (**E**) RT-qPCR validation of three target genes across four technical replicates per group: *BRD1* (a down-regulated gene on chr 22); *RIC3* (an up-regulated gene on chr 11); and *CLIC5* (a down-regulated gene on chr 6). A Student’s t-test was used to test delta CT values significant differences. (**F**) CAMERA cell type enrichment of under-expressed DEGs (x-axis) according to seven immune cell types (y-axis) reveals strong enrichment of CD56+ genes. (**G**) CIBERSORTx cell type predictions reveal a significant reduction in the frequency CD56+ cells among Class II mutations. (**H**) CyTOF validates estimated cell type proportions on a subset of controls and participants with Class II mutations. Scaled frequencies across all participants for major and minor immune cell populations are presented in heatmap form (right). Boxplots of the two immune populations with significant differences (*p*<0.05, linear model) associated with Class II mutations (left).

Of the significantly under-expressed genes associated with Class II mutations, 31 genes were located on 22q13.3 while the remaining (∼82%) were not linked to this genomic region (**Figure 2B**). We identified genes chloride intracellular channel 5 (*CLIC5*) and keratin type I cytoskeletal 23 (*KRT23*) to be among the most significant under-expressed genes in Class II mutations not located on 22q13.3. We also identified RIC3 acetylcholine receptor chaperone (*RIC3*) and paternally expressed 3 (*PEG3*) to be among the most significant over-expressed genes in Class II mutations. A competitive gene-set ranking approach was used to functionally annotate Class II-related genes, revealing over-expressed genes enriched for processes related to nonsense mediated decay (FDR *p*=9.3×10^−16^), protein translation (FDR *p*=4.0×10^−14^) and cell cycle check points (FDR *p*=0.003), while under-expressed genes enriched for extracellular matrix organization (FDR *p*=0.0001), NCAM1 (also known as CD56) interactions (FDR *p*=0.004), voltage gated calcium channel activity (FDR *p*=0.006), and glycosphingolipid metabolism (FDR *p*=0.03) (**Figure 2C-D**), among other processes (**Table S2**). To ensure confidence of our results, we performed technical validation of three genes of interest by RT-qPCR, which confirmed significant under-expression of *BRD1* and *CLIC4*, as well as significant over-expression of *RIC3* in Class II participants relative to Class I mutations and unaffected controls (**Figure 2E**).

To support these functional enrichment observations, we tested whether the candidate dysregulated genes indeed interact with each other at the protein level. A significant overrepresentation of direct protein-protein interactions (PPI) was identified among differentially expressed genes in PMS participants with Class II mutations (*p*<1.0e-16, observed edges=228, expected edges=112) (**Figure S4**). As expected, disrupted genes on 22q13.3 displayed a higher average number of interactions (average node degree=2.32) relative to under-expressed and over-expressed genes in PMS participants with Class II mutations (average node degree=1.77 and 0.50, respectively). The peripheral blood PPI network derived from participants Class II mutations was again enriched for components related to NCAM1 signaling and cytotoxic immune cell signatures, and featured several under-expressed hub genes, including *NCAM1*, perforin 1 (*PRF1*), and interleukin 2 receptor subunit beta (*IL2RB*), as well as genes T-Box transcription factor 21 (*TBX21*) and sphingosine-1-phosphate receptor 5 (*S1PR5*), which are critical for the maturation and recruitment of CD56+ natural killer (NK) cells into the periphery^35,36^.

### Predicting reduced NK cell-specific expression and cellular proportions in Class II mutations

A multi-step approach explored the cellular origins of the differentially expressed genes in Class II mutations. First, we collected genes that are significantly and highly expressed across seven main immune cell types leveraging an existing scRNA-seq experiment (*see Methods*). Using the same gene-set ranking approach as above, we performed cell type enrichment analysis and identified a significant enrichment of CD56+ NK cell genes among under-expressed genes in Class II mutations (FDR *p*=6.8×10^−10^) (**Figure 2F**). Notably, we also observed an enrichment for CD56+ NK cell genes among nominally significant under-expressed genes in participants Class I mutations (FDR *p*=1.5×10^−5^) (**Figure S5A-B**). Second, we performed the reverse approach by querying the expression of the 208 under-expressed genes in Class II mutations within thousands of single peripheral blood mononuclear cells across three independent experiments (*see Methods*). These analyses confirm that under-expressed genes associated Class II mutations are consistently and highly expressed in CD56+ NK cells (**Figure S6**). Third, we performed cell type deconvolution analysis of the bulk peripheral blood transcriptome data using an independent cell type-specific reference marker list, and confirmed a significant reduction in the proportion of estimated CD56+ NK cells in Class II mutations compared to unaffected controls (*p*=0.007) (**Figure 2G**). Notably, a general reduction in the proportion of CD56+ NK cells was also observed among Class I mutations, albeit non-significant (*p*=0.104). Fourth, we re-computed our differential gene expression analyses for i) all PMS participants and ii) Class II mutations relative to unaffected controls covarying for CD56+ NK cellular proportions. Adjusting for CD56+ NK cells had the largest effect on differential gene expression, and removed ∼69% of differentially expressed genes in Class II mutations relative to unaffected controls (**Figure S5C**). Fifth, we identified 25 genes, including *S1PR5*, that were highly expressed in CD56+ NK cells via scRNA-seq that were also significantly under-expressed among Class II mutations and performed unsupervised hierarchical clustering, which accurately classified 82% (29/35) of participants with Class II deletion from remaining samples (**Figure S5D**). Finally, given the critical role of S1PR5 to recruit NK cells into the peripheral circulation and to bind lipid signaling molecule sphingosine 1-phosphate (S1P)^35,36^, we asked which of the disrupted genes on 22q13.3 might play a role in regulating *S1PR5* expression and/or sphingolipid metabolism. While the vast majority the 52 blood expressed genes on 22q13.3 were highly correlated with *S1PR5*, we found that the expression of ceramide kinase (*CERK*), and parsing Class II mutations according to those with the disruption of CERK relative to the remainder of Class II mutations, was moderately predictive of *S1PR5* expression levels (**Figure S7**). This observation was strengthened by the direct PPIs observed between CERK and SIPR5 (**Figure S4**).

### Mass cytometry validates reduced peripheral CD56+ NK cells and egress to the periphery in Class II mutations

To validate these predictions, we performed cytometry by time-of-flight (CyTOF)-based immunophenotyping on a subset of PMS participants with Class II mutations (*n*=5) and an age and sex matched control group (*n*=4). While both controls and Class II mutations had similar distributions of major immune cell subsets in peripheral blood, the frequencies of finer immune cell types were significantly altered (**Figure 2H**). Specifically, we observed a significant increase in the proportions of CD3+ CD4-CD8- (double-negative) T cells and a significant reduction in the proportions of CD56+ CD16-NK cells in Class II mutations (*p*=0.001, *p*=0.03, respectively) (**Figure 2H**), validating our *in silico* predictions. Collectively, these results indicate that Class II mutations are associated with unique peripheral blood transcriptional changes, which might be explained by alterations in the underlying cellular composition of CD56+ NK cells and/or related cell-specific gene expression programs.

### Secondary exploratory analysis reveals transcriptomic predictors of ABC-SW

A secondary exploratory analysis examined relationships between collected clinical phenotypes and peripheral blood gene expression across all 68 PMS participants. While few significant associations (*n*=3) were observed at a *FDR* < 5%, relaxing our statistical assumption of significance to a *FDR* < 10% uncovered 1,017 genes, which were predominately associated with differences in Aberrant Behavior Checklist Social Withdrawal subscale^37^ (ABC-SW) scores (*n*=1,013 genes) (**Figure S8A-B, Table S3**). Genes positively associated with ABC-SW were significantly enriched for RNA binding, splicing and protein translation, while negatively associated genes were implicated in transcription coregulatory activity, chromatin organization, and histone modifications (**Figure S8C, Table S3**). Genes negatively associated with ABC-SW were also significantly enriched for genes that implicate genetic risk for intellectual disability, ASD, developmental delay, and educational attainment, as well as differentially expressed genes in postmortem brain tissue from individuals with ASD (**Figure S8D-E**). Notably, ABC-SW-related genes were not enriched for an immune cell type signature nor were associated with differences in estimated cell type proportions (**Figure S8A**).

### Class II mutations, but not Class I mutations, reduce sphingolipid metabolism

Global metabolomic data were generated across a subset of 54 participants, comprised of Class I mutations (*n*=11), Class II mutations (*n*=14), and an age and sex matched control group (*n*=29), half of which were familial related unaffected siblings (∼51%). Global metabolomic profiling identified 1,045 high confidence metabolites across all 54 participants, which were largely made of lipids (37%), amino acids (19%), xenobiotics (13%), an unknown category (15%) and six other less frequent super pathways (**Figure 3A**). We modelled for differential changes in metabolite abundance in PMS participants as described above, and identified 10 metabolites significantly associated with all PMS participants, 9 metabolites associated with Class I mutations, and 24 metabolites associated with Class II mutations relative to unaffected controls (**Figure 3A Table S4**). Notably, the pattern of metabolomic effect sizes observed for Class II mutations was consistent with transcriptome-wide effect sizes (**Figure 3B, Figure 2B**), in that the majority of metabolites were less abundant in participants with Class II mutations. Of the 24 altered metabolites associated with Class II mutations, 21 were less abundant relative to unaffected controls, including 10 metabolites catalogued as part the of the sphingomyelin lipid family (**Figure S9**). These findings also provide validation the observed transcriptomic alterations of reduced expression of genes enriched for glycosphingolipid metabolism (**Figure 2**). Unsupervised hierarchical clustering of these 24 metabolites distinguished 85% (12/14) of participants with Class II mutations from the remaining samples (**Figure 4C**). Finally, we performed an integrated analysis of differentially expressed genes and differentially abundant metabolites to elucidate their combined effect on key metabolomic pathways in participants with Class II mutations. This analysis confirmed significant changes in sphingolipid metabolism followed by alterations in arginine and proline metabolism and linoleic acid metabolism (**Figure 4D, Table S4**).

**Figure 3.**
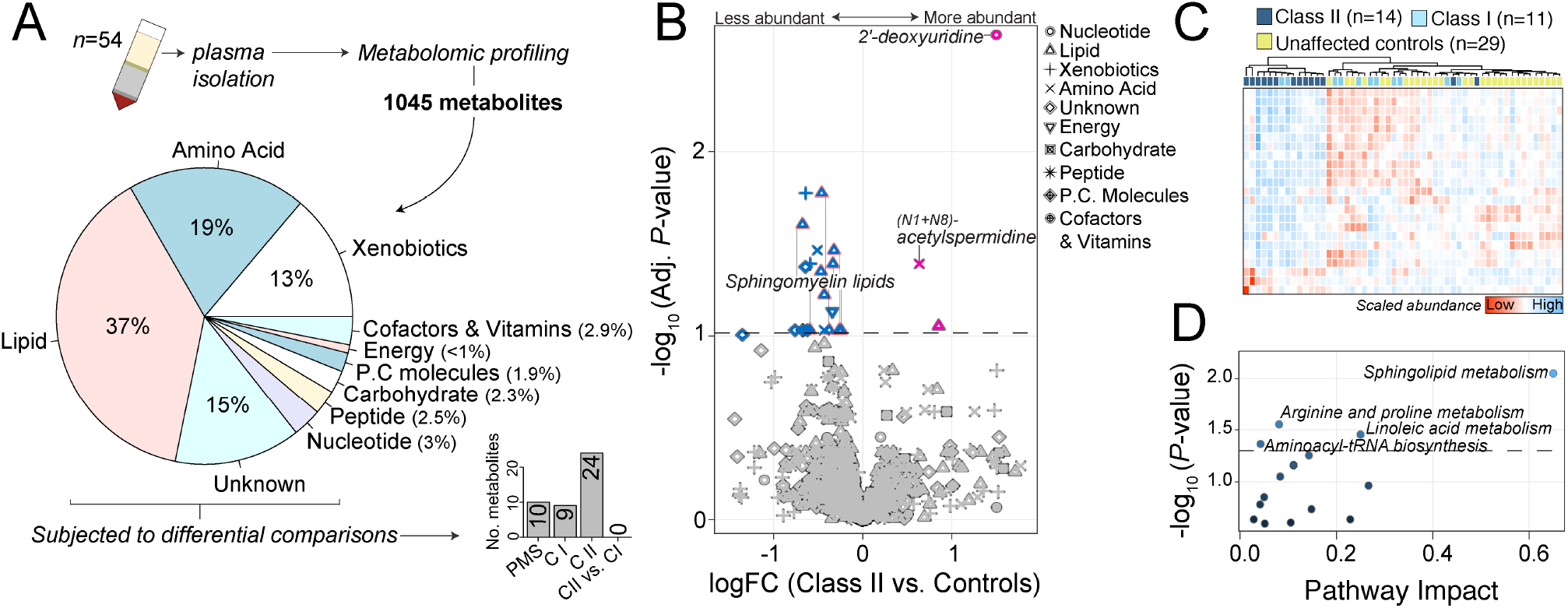
Plasma metabolomic profiling and alterations in participants with Class II mutations. (**A**) Top inset: Plasma was collected from 54 participants and subjected to unbiased metabolomic profiling, which generated 1045 high-confidence metabolites for subsequent analysis. The majority of detected metabolites classified as either lipids (37%), amino acids (19%), xenobiotics (13%), unknown (15%) or six other less frequent categories. Bottom inset: Differential abundance of metabolites was tested and the number of significant metabolites for each comparison are displayed. (**B**) Volcano plot of class II differentially abundant metabolites (DAMs) relative to unaffected controls depicting log fold-change (logFC; x-axis) and –log_10_ FDR adjusted p-value. The dotted horizontal line indicates a cut-off of FDR < 0.1. Metabolites are uniquely shaped according to ten super pathway categories and colored by more (red) or less (blue) abundant in participants with Class II mutations. Ten sphingomyelin metabolites are outlined in pink borders. (**C**) Unsupervised clustering of 24 metabolites significantly altered by Class II mutations correctly classify 85% (n=12) of Class II mutations from the remaining samples. Heatmap depicts high (red) and low (blue) relative scaled abundance for each metabolite. (**D**) Pathway analysis of metabolites altered in Class II mutations reveals significant pathway enrichment (y-axis; -log_10_ p-value) for spingolipid metabolism and three other metabolism pathways relative to pathway impact (x-axis). Pathway impact is a combination of the centrality and pathway enrichment results computed by adding the importance measures of each of matched metabolite and dividing by the sum of the importance measures of all metabolites in each pathway.

## DISCUSSION

While increased frequency and severity of PMS phenotypes are associated with larger deletions, the molecular perturbations that result from specific genetic alteration remain poorly understood. The current study presents the largest set of PMS genotypes associated with peripheral blood gene expression and global serum metabolites conducted to date, and highlights several candidate genes, pathways, metabolites and cell types uniquely linked to PMS cases with Class II mutations, despite the disruption of *SHANK3* in all participants. This suggests that SHANK3 alone is not responsible for the molecular alterations observed in the peripheral circulation. Specifically, these findings reveal that PMS participants with Class II mutations display decreased expression of key cytotoxic immune cell signatures and related processes, reductions in the proportions of cytotoxic cell types, and reduced sphingolipid metabolism. Below we discuss the biological and clinical implications of our results.

Of the disrupted genes in the 22q13.3 region, 52 genes (∼40%) were detected in peripheral blood, reduced in expression patterns, and largely predictive of deletion size, classifying 30/35 PMS participants with Class II mutations from all other participants by unsupervised hierarchical clustering (**Figure 1**). Many of these disrupted, under-expressed genes are individually linked to independent rare disorders and are known to partake in diverse cellular signaling systems, including inflammatory responses (*e*.*g. MAPK11*)^38^, glycosylation (*e*.*g. ALG12*)^39^, mitochondrial translation (*e*.*g. TRMU*)^40^, kinase activity (*e*.*g. PARVB, PARVG, CERK, PIM3*)^41-43^, tubulin ligase activity (*e*.*g. TTLL12, TTL1*)^44^, histone acetyltransferase activity (*e*.*g. BRD1*)^45,46^, and sphingolipid metabolism (*e*.*g. ARSA, CERK*)^47,48^ among others. For example, under-expression of modulator of VRAC current 1 (*MLC1*) and peroxisome proliferator activated receptor alpha (*PPARA*) were also observed in Class II mutations, and these genes are linked to megalencephalic leukoencephalopathy with subcortical cysts disease and are known to interact with several ion channels/transporters and accessory proteins^49,50^. While these genes were also expressed across a broad collection of other human tissues (**Figure S2**), our results highlight the utility of peripheral blood transcriptome profiling as an accessible, alternative diagnostic read out to validate genotypic variation on 22q13.3 for PMS and several other monogenic disorders with a locus on 22q13.3.

Beyond alterations of disrupted genes on 22q13.3, the most significant transcriptomic changes were for those under-expressed genes in PMS participants with Class II mutations, related to CD56+ NK cell signatures. Analysis by CyTOF confirmed a reduction in CD56+ CD16-cells in Class II mutations, which are less cytotoxic than CD56+ CD16+ cells but produce greater amounts of cytokines in response to environmental cues. Under-expression of sphingosine-1-phosphate receptor 5 (*S1PR5*) was among one of many NK cell-related genes that was under-expressed in Class II mutations (**Figure 2, Figure S4-5**), and this gene is well-known to promote recirculation and recruitment of CD56+ NK cells into the periphery^35,36,51^. NK cells express *S1PR5*, and in mice deficient in this receptor, NK cell distribution was altered, with reduced NK cell numbers in blood and spleen and increased numbers in the lymph node and bone marrow^52,53^. We also observed decreased expression of T-Box Transcription Factor 21 (*TBX21*) in PMS participants with Class II mutations, which is required for the final maturation of NK cells^54^ and is also known to induce *S1PR5*^35,36^, supporting a mechanism whereby reduced *TBX21* lends to lower expression of *S1PR5* and ultimately reduced CD56+ NK cell proportions (**Figure 2G**), which may increase susceptibility to viral infections in these individuals. While the rate of viral infections has not been deeply studied in PMS, the largest PMS phenotypic study to date describes that 38% of PMS participants with Class I mutations (29/76) and 51% of those with Class II mutations (44/87) report recurrent infections^8^, supportive of a smaller independent study where larger deletion sizes were associated with increased frequency of recurrent infections^10^. Notably, there are at least 46 single gene primary immunodeficiencies that feature NK cell deficiencies^55^, defined as either the absence of NK cells and their functions or the presences of defective NK cells within peripheral blood lymphocytes. While various therapeutics have been applied to treat individuals with NK cell deficiencies, most approaches have focused on treating susceptibility to viral infections via the application of prophylactic antiviral drugs. Nevertheless, anecdotal cases have described apparent success using acyclovir, ganciclovir and cytokine therapies^56-60^, such as IFN-α, to induce NK cell cytotoxic functions. However, these approaches require further investigation and consideration is currently done on a case-by-case basis.

In an effort to resolve which of the disrupted genes in the extended 22q13.3 region might have the most direct effect on cytotoxic cell recruitment and/or sphingolipid metabolism, we predicted that the disrupted gene ceramide kinase (*CERK*), may also play a role (**Figure S6**). CERK is required for the phosphorylation of ceramide, which is the centerpiece of the sphingolipid metabolism and generates ceramide 1-phosphate (C1P)^48,61^. Both CERK and C1P have been implicated in cellular proliferation, apoptosis and inflammation^62,63^. Our findings from serum metabolomic profiling support these results by directly implicating reductions of sphingolipid metabolism in PMS participants with Class II mutations (**Figure 3**). In addition to phosphorylation by CERK, ceramide can be hydrolysed to sphingosine, which is phosphorylated to sphingosine-1-phosphate (S1P) by sphingosine kinases^47,48,61^. Both C1P and S1P are bioactive molecules critical for immune function and inflammation^47,48,61-63^, but also play an important role in neurotransmitter release and synaptic transmission in the brain^64^. Whereas ceramide and sphingosine are associated with cellular growth arrest and apoptosis, S1P is associated with cellular survival and suppression of apoptosis^61-64^. To this end, we anticipate that reduced sphingolipid metabolism in PMS participants with Class II mutations is associated with down-regulation of ceramide biosynthesis and/or S1P synthesis. Notably, S1P signaling via S1PR5 is particularly important for regulating NK cell migration and cytotoxicity and gradients of S1P drive NK cell chemotaxis^35,36,65,66^, essential for the mobilization of NK cells to inflamed organs, supporting our transcriptomic results. Taken together, our data provide preliminary evidence for a mechanistic model linking large 22q13.3 deletions to reductions in NK cell related gene expression signatures (*TBX21, S1PR5, NCAM1* and partners) and CD56+ cellular proportions in the periphery, as well as reductions in sphingolipid metabolism, which would otherwise lend to the recruitment and survival of these cells in the peripheral circulation. Notably, alterations of lipid metabolism, including S1P, have been reported in serum and postmortem brain tissues from individuals with ASD^67,68^, suggesting that impairment of lipid metabolism pathways may contribute to the pathology of ASD more broadly. Several therapeutic agents have been developed to modulate sphingolipid metabolism, including stress-signalling molecules tumour necrosis factor (TNF)-α and interleukin-1β (IL-1β), to induce activation of sphingomyelinases^69,70^, which can also increase ceramide and subsequent ceramide-dependent responses (*i*.*e*. cell death and/or arrest).

Our secondary exploratory analysis also identified several genes that are both positively and negatively associated with variations in ABC-SW across all PMS cases (FDR < 10%) (**Figure S7**). Notably, genes negatively associated with ABC-SW were implicated in transcription coregulatory activity, chromatin organization, histone modifications, and were significantly enriched for genes that implicate genetic risk for neurodevelopmental disorders. Thus, individuals with high ABC-SW scores display reduced levels expression for these genes. Given the interest in the ABC-SW as a clinical outcome assessment of treatment efficacy in clinical trials of PMS^71,72^, peripheral biomarkers that scale ABC-SW severity may serve as a valuable resource to monitor treatment responses and outcomes in PMS and other disorders that present with social withdrawal phenotypes. However, further follow-up of these genes and their dynamic expression profiles following administration of such therapeutic agents is warranted.

Our study does present some limitations. First, in the current report, a clinician-made assessment was used to characterize ‘recurrent infections’ in PMS participants, defined as more than two pneumonia or sinus infections per year. Under this measure, ∼54% of PMS participants with Class II mutations (13/25) and ∼50% of those with Class I mutations (11/22) report recurrent infections (**Table 1**). This broad definition does not encompass viral infections, nor specifically delineate the types of observed infections, severities, annual frequencies nor medication(s) described; thus, limiting our ability to causally link 22q13.3 deletion sizes and the reported transcriptomic and metabolomic alterations with specific immune phenotypes. Nevertheless, in addition to immune function, NK cells and NCAM1 are also present in the human brain and are implicated in several brain-related mechanisms, including neuronal migration, synaptic plasticity and clearance of α-syn aggregates through the lysosomal pathway^73-75^. Thus, more work is needed to fully dissect the relationship between the molecular and clinical expressivity observed in PMS. Second, the subset of individuals with serum metabolomics profiling (*n*^*total*^=54) was predominately independent from those with peripheral blood transcriptomic data (*n*^*total*^=92), with 29 overlapping individuals. While this variation can increase synergy and confidence of the reported alterations across transcriptomic and metabolomics datasets, increasing the availability of paired data will better power discovery and interpretation of the reported cytotoxic cell signatures and sphingolipid metabolomics changes in PMS. Finally, while sphingolipids are well known mediators of cell fate, they may also change in response to drug treatment, and such alterations might also reflect differential responses to existing treatment regimens.

## CONCLUSION

Taken together, we show that participants with Class II mutations present significant peripheral transcriptomic and metabolomics alterations implicating reductions in cytotoxic immune cell signatures, CD56+ CD16-cell proportions, and sphingolipid metabolism, which may contribute to a more the severe and variable phenotype in PMS. More broadly, this work demonstrates the utility of studying molecules in the peripheral blood of individuals with PMS, which is a readily available specimen type in clinical practice. It is worth noting that this combination of data is not expected to successfully shed light on disrupted genes and pathways if the affected region(s) is not expressed in the analyzed tissue or if the effects of the causal variants do not affect the expression of the gene. Therefore, expert evaluation is required when prioritizing candidate genes using RNA-seq data. We can expect that combining information from multiple ‘omics’ sources will only further improve diagnosis and define molecular subtypes of PMS and other rare disease cases in the future.

## Data Availability

All data produced in the present study are available upon reasonable request to the authors

## LIST OF ABBREVIATIONS

PMS: Phelan-McDermid syndrome
ASD: Autism spectrum disorder
SHANK3: SH3 And Multiple Ankyrin Repeat Domains 3
Class I mutations: sequence variants or small deletions on 22q13.3, including *SHANK3* only or *SHANK3* with *ARSA* and/or *ACR* and *RABL2B*
Class II mutations: larger 22q13.3 deletions relative to Class I mutations
DSC: Developmental Synaptopathies Consortium
ABC-SW: ABC-social withdrawal
NK cells: Natural killer cells

## DECLARATIONS

### Ethics approval and consent to participate

This study was approved by the Institutional Review Board (IRB) for the protection of human subjects at Mount Sinai (Study IDs: 98–0436, 10-0527, 12-1718) and Boston Children’s Hospital (Study ID: P00013300), which serves as the central IRB for the Developmental Synaptopathies Consortium (DSC).

### Consent for publication

Not applicable

### Competing interests

A.K. receives research support from AMO Pharma and consults to Acadia, Alkermes, Neuren and GW Pharma. He serves on Scientific Advisory Boards for Ovid Therapeutics, Jaguar Therapeutics and Ritrova Therapeutics. M.S. reports grant support from Novartis, Biogen, Astellas, Aeovian, Bridgebio and Aucta. He has served on Scientific Advisory Boards for Novartis, Roche, Regenxbio and Alkermes. The remaining authors declare that they have no competing interests.

### Availability of data and materials

Original RNA-sequencing FASTQ files are available at the Gene Expression Omnibus under accession number GSEXXXXX. Original CyTOF FCS files are available at Immport under accession number SDYXXXX (https://www.immport.org). Global metabolomic data are available in Supplemental Table 4. (accession numbers will be updated prior-to paper acceptance).

### Funding

The Developmental Synaptopathies Consortium (U54NS092090) is part of the Rare Diseases Clinical Research Network (RDCRN), an initiative of the Office of Rare Diseases Research (ORDR), National Center For Advancing Translational Sciences (NCATS). Research reported in this publication was supported by the National Institute Of Neurological Disorders And Stroke of the National Institutes of Health (NINDS), Eunice Kennedy Shriver National Institute Of Child Health & Human Development (NICHD), National Institute Of Mental Health (NIMH) and National Center For Advancing Translational Sciences (NCATS). The content is solely the responsibility of the authors and does not necessarily represent the official views of the National Institutes of Health (NIH).

### Author contributions

*Concept and design*: JDB, MSB. *Clinical evaluations, sample ascertainment, clinical data curation*: TS, BC, MS, EBK, LS, AT, CMP, JAB, AK, JDB. *RNA extraction and bio-specimen handling*: MSB. *Data analysis*: MSB, XF, RP, AO. *Data interpretation*: MSB, AST, AK, JDB. All authors contributed to writing, reading and approved the final manuscript.

## Acknowledgements

We are sincerely indebted to the generosity of the families and patients in PMS clinics across the United States who contributed their time and effort to this study. We would also like to thank the Phelan-McDermid Syndrome Foundation for their continued support in PMS research.

## Corresponding authors

Correspondence to Michael S. Breen and Joseph D. Buxbaum

## TABLES TITLES AND LEGENDS

**Supplemental Table 1. Sample level clinical and medical phenotypes used in the current study**.

**Supplemental Table 2. Summary statistics for differential gene expression, pathway and protein interaction analyses**.

**Supplemental Table 3. Exploratory analysis of gene expression associated with ABC-SW**.

**Supplemental Table 4. Metabolomic data and differential comparison summary statistics**.

## Notes

### Author Declarations

Ethics committee/IRB of the Icahn School of Medicine at Mount Sinai and Boston Childrens Hospital gave ethical approval for this work.

## REFERENCES

1. Betancur C, Buxbaum JD. SHANK3 haploinsufficiency: a “common” but underdiagnosed highly penetrant monogenic cause of autism spectrum disorders. Mol Autism. 2013;4(1):17.

2. Leblond CS, Nava C, Polge A, Gauthier J, Huguet G, Lumbroso S, Giuliano F, Stordeur C, Depienne C, Mouzat K, et al. Meta-analysis of SHANK mutations in autism spectrum disorders: a gradient of severity in cognitive impairments. PLoS Genet. 2014;10(9):e1004580.

3. Boccuto L, Lauri M, Sarasua SM, Skinner CD, Buccella D, Dwivedi A, Orteschi D, Collins JS, Zollino M, Visconti P, et al. Prevalence of SHANK3 variants in patients with different subtypes of autism spectrum disorders. Eur J Hum Genet. 2013;21(3):310–6.

4. De Rubeis S, Siper PM, Durkin A, Weissman J, Muratet F, Halpern D, Trelles MDP, Frank Y, Lozano R, Wang AT, et al. Delineation of the genetic and clinical spectrum of Phelan-McDermid syndrome caused by SHANK3 point mutations. Mol Autism. 2018;9:31.

5. Mitz AR, Philyaw TJ, Boccuto L, Shcheglovitov A, Sarasua SM, Kaufmann WE, Thurm A. Identification of 22q13 genes most likely to contribute to Phelan McDermid syndrome. Eur J Hum Genet. 2018;26(3):293–302.

6. Soorya L, Kolevzon A, Zweifach J, Lim T, Dobry Y, Schwartz L, Frank Y, Wang AT, Cai G, Parkhomenko E, et al. Prospective investigation of autism and genotype-phenotype correlations in 22q13 deletion syndrome and SHANK3 deficiency. Mol Autism. 2013;4(1):18.

7. Sarasua SM, Dwivedi A, Boccuto L, Chen CF, Sharp JL, Rollins JD, Collins JS, Rogers RC, Phelan K, DuPont BR. 22q13.2q13.32 genomic regions associated with severity of speech delay, developmental delay, and physical features in Phelan-McDermid syndrome. Genet Med. 2014;16(4):318–28.

8. Levy T, Foss-Feig JH, Betancur C, Siper PM, Trelles-Thorne MD, Halpern D, Frank Y, Lozano R, Layton C, Britvan B, Bernstein JA. Strong evidence for genotype–phenotype correlations in Phelan-McDermid syndrome: results from the developmental synaptopathies consortium. Human Molecular Genetics. 2021 Sep 24.

9. Bonaglia MC, Giorda R, Beri S, De Agostini C, Novara F, Fichera M, et al. Molecular mechanisms generating and stabilizing terminal 22q13 deletions in 44 subjects with Phelan/McDermid syndrome. PLoS Genet. 2011;7:e1002173.

10. Wilson HL, Wong AC, Shaw SR, Tse WY, Stapleton GA, Phelan MC, Hu S, Marshall J, McDermid HE. Molecular characterisation of the 22q13 deletion syndrome supports the role of haploinsufficiency of SHANK3/PROSAP2 in the major neurological symptoms. Journal of medical genetics. 2003 Aug 1;40(8):575–84.

11. Durand CM, Perroy J, Loll F, Perrais D, Fagni L, Bourgeron T, Montcouquiol M, Sans N. SHANK3 mutations identified in autism lead to modification of dendritic spine morphology via an actin-dependent mechanism. Mol Psychiatry. 2012;17(1):71–84.

12. Halbedl S, Schoen M, Feiler MS, Boeckers TM, Schmeisser MJ. Shank3 is localized in axons and presynaptic specializations of developing hippocampal neurons and involved in the modulation of NMDA receptor levels at axon terminals. J Neurochem. 2016;137(1):26–32.

13. Lee K, Vyas Y, Garner CC, Montgomery JM. Autism-associated Shank3 mutations alter mGluR expression and mGluR-dependent but not NMDA receptor-dependent long-term depression. Synapse. 2019;73(8):e22097.

14. Mitz AR, Philyaw TJ, Boccuto L, Shcheglovitov A, Sarasua SM, Kaufmann WE, Thurm A. Identification of 22q13 genes most likely to contribute to Phelan McDermid syndrome. European Journal of Human Genetics. 2018 Mar;26(3):293–302.

15. Frye RE, Cox D, Slattery J, Tippett M, Kahler S, Granpeesheh D, Damle S, Legido A, Goldenthal MJ. Mitochondrial dysfunction may explain symptom variation in Phelan-McDermid syndrome. Scientific reports. 2016 Jan 29;6(1):1–2.

16. Schenkel LC, Aref-Eshghi E, Rooney K, Kerkhof J, Levy MA, McConkey H, Rogers RC, Phelan K, Sarasua SM, Jain L, Pauly R. DNA methylation epi-signature is associated with two molecularly and phenotypically distinct clinical subtypes of Phelan-McDermid syndrome. Clinical Epigenetics. 2021 Dec;13(1):1–7.

17. Jain L, Oberman LM, Beamer L, Cascio L, May M, Srikanth S, Skinner C, Jones K, Allen B, Rogers C, Phelan K. Genetic and metabolic profiling of individuals with Phelan-McDermid syndrome presenting with seizures. Clinical Genetics. 2021 Oct 19.

18. Voineagu, I., Wang, X., Johnston, P., Lowe, J.K., Tian, Y., Horvath, S., Mill, J., Cantor, R.M., Blencowe, B.J. and Geschwind, D.H., 2011. Transcriptomic analysis of autistic brain reveals convergent molecular pathology. Nature, 474(7351), pp.380–384.

19. Gupta S, Ellis SE, Ashar FN, Moes A, Bader JS, Zhan J, West AB, Arking DE. Transcriptome analysis reveals dysregulation of innate immune response genes and neuronal activity-dependent genes in autism. Nature communications. 2014 Dec 10;5(1):1–8.

20. Tylee, D.S., Hess, J.L., Quinn, T.P., Barve, R., Huang, H., Zhang-James, Y., Chang, J., Stamova, B.S., Sharp, F.R., Hertz-Picciotto, I. and Faraone, S.V., 2017. Blood transcriptomic comparison of individuals with and without autism spectrum disorder: A combined-samples mega-analysis. American Journal of Medical Genetics Part B: Neuropsychiatric Genetics, 174(3), pp.181–201.

21. Frésard L, Smail C, Ferraro NM, Teran NA, Li X, Smith KS, Bonner D, Kernohan KD, Marwaha S, Zappala Z, Balliu B. Identification of rare-disease genes using blood transcriptome sequencing and large control cohorts. Nature medicine. 2019 Jun;25(6):911–9.

22. Signorelli M, Ebrahimpoor M, Veth O, Hettne K, Verwey N, García-Rodríguez R, Tanganyika-deWinter CL, Lopez Hernandez LB, Escobar Cedillo R, Gómez Díaz B, Magnusson OT. Peripheral blood transcriptome profiling enables monitoring disease progression in dystrophic mice and patients. EMBO Molecular Medicine. 2021 Apr 9;13(4):e13328.

23. Breen MS, Stein DJ, Baldwin DS. Systematic review of blood transcriptome profiling in neuropsychiatric disorders: guidelines for biomarker discovery. Human Psychopharmacology: Clinical and Experimental. 2016 Sep;31(5):373–81.

24. Dobin A, Davis CA, Schlesinger F, Drenkow J, Zaleski C, Jha S, Batut P, Chaisson M, Gingeras TR. STAR: ultrafast universal RNA-seq aligner. Bioinformatics. 2013;29(1):15–21.

25. Liao Y, Smyth GK, Shi W. featureCounts: an efficient general purpose program for assigning sequence reads to genomic features. Bioinformatics. 2014;30(7):923–30

26. Ritchie ME, Phipson B, Wu D, Hu Y, Law CW, Shi W. Smyth GK: limma powers differential expression analyses for RNA-sequencing and microarray studies. Nucleic Acids Res. 2015;43(7):e47.

27. Wu D, Smyth GK. Camera: a competitive gene set test accounting for inter-gene correlation. Nucleic acids research. 2012 Sep 1;40(17):e133..

28. Zheng GX, Terry JM, Belgrader P, Ryvkin P, Bent ZW, Wilson R, Ziraldo SB, Wheeler TD, McDermott GP, Zhu J, Gregory MT. Massively parallel digital transcriptional profiling of single cells. Nature communications. 2017 Jan 16;8(1):1–2.

29. Satija R, Farrell JA, Gennert D, Schier AF, Regev A. Spatial reconstruction of single-cell gene expression data. Nature biotechnology. 2015 May;33(5):495–502.

30. Steen CB, Liu CL, Alizadeh AA, Newman AM. Profiling cell type abundance and expression in bulk tissues with CIBERSORTx. InStem Cell Transcriptional Networks 2020 (pp. 135–157). Humana, New York, NY.

31. Evans, A. M., DeHaven, C. D., Barrett, T., Mitchell, M. & Milgram, E. Integrated, nontargeted ultrahigh performance liquid chromatography/electrospray ionization tandem mass spectrometry platform for the identification and relative quantification of the small-molecule complement of biological systems. Anal. Chem. 81, 6656–6667 (2009).R

32. Dehaven, C. D., Evans, A. M., Dai, H. & Lawton, K. A. Organization of GC/MS and LC/MS metabolomics data into chemical libraries. J. Cheminform. 2, 9 (2010).

33. Pang Z, Chong J, Zhou G, de Lima Morais DA, Chang L, Barrette M, Gauthier C, Jacques PÉ, Li S, Xia J. MetaboAnalyst 5.0: narrowing the gap between raw spectra and functional insights. Nucleic acids research. 2021 May 21.

34. Wishart DS, Tzur D, Knox C, Eisner R, Guo AC, Young N, Cheng D, Jewell K, Arndt D, Sawhney S, Fung C. HMDB: the human metabolome database. Nucleic acids research. 2007 Jan 1;35(Suppl_1):D521–6.

35. Jenne CN, Enders A, Rivera R, Watson SR, Bankovich AJ, Pereira JP, Xu Y, Roots CM, Beilke JN, Banerjee A, Reiner SL. T-bet–dependent S1P5 expression in NK cells promotes egress from lymph nodes and bone marrow. Journal of Experimental Medicine. 2009 Oct 26;206(11):2469–81.

36. Evrard M, Wynne-Jones E, Peng C, Kato Y, Christo SN, Fonseca R, Park SL, Burn TN, Osman M, Devi S, Chun J. Sphingosine 1-phosphate receptor 5 (S1PR5) regulates the peripheral retention of tissue-resident lymphocytes. Journal of Experimental Medicine. 2022 Jan 3;219(1).

37. Aman MG, Singh NN, Stewart AW, Field CJ. Psychometric characteristics of the aberrant behavior checklist. American journal of mental deficiency. 1985 Mar.

38. Arthur JS, Ley SC. Mitogen-activated protein kinases in innate immunity. Nature Reviews Immunology. 2013 Sep;13(9):679–92.

39. Grubenmann CE, Frank CG, Kjaergaard S, Berger EG, Aebi M, Hennet T. ALG12 mannosyltransferase defect in congenital disorder of glycosylation type lg. Human molecular genetics. 2002 Sep 15;11(19):2331–9.

40. Zeharia A, Shaag A, Pappo O, Mager-Heckel AM, Saada A, Beinat M, Karicheva O, Mandel H, Ofek N, Segel R, Marom D. Acute infantile liver failure due to mutations in the TRMU gene. The American Journal of Human Genetics. 2009 Sep 11;85(3):401–7.

41. Yen, C.F., Wang, H.S., Lee, C.L. and Liao, S.K., 2014. Roles of integrin-linked kinase in cell signaling and its perspectives as a therapeutic target. Gynecology and Minimally Invasive Therapy, 3(3), pp.67–72.

42. Sugiura M, Kono K, Liu H, Shimizugawa T, Minekura H, Spiegel S, Kohama T. Ceramide kinase, a novel lipid kinase: molecular cloning and functional characterization. Journal of Biological Chemistry. 2002 Jun 28;277(26):23294–300.

43. Beharry Z, Mahajan S, Zemskova M, Lin YW, Tholanikunnel BG, Xia Z, Smith CD, Kraft AS. The Pim protein kinases regulate energy metabolism and cell growth. Proceedings of the National Academy of Sciences. 2011 Jan 11;108(2):528–33.

44. Brants J, Semenchenko K, Wasylyk C, Robert A, Carles A, Zambrano A, Pradeau-Aubreton K, Birck C, Schalken JA, Poch O, de Mey J. Tubulin tyrosine ligase like 12, a TTLL family member with SET- and TTL-like domains and roles in histone and tubulin modifications and mitosis. PloS one. 2012 Dec 12;7(12):e51258.

45. Christensen JH, Elfving B, Müller HK, Fryland T, Nyegaard M, Corydon TJ, Nielsen AL, Mors O, Wegener G, Børglum AD. The Schizophrenia and Bipolar Disorder associated BRD1 gene is regulated upon chronic restraint stress. European Neuropsychopharmacology. 2012 Sep 1;22(9):651–6.

46. Fryland T, Christensen JH, Pallesen J, Mattheisen M, Palmfeldt J, Bak M, Grove J, Demontis D, Blechingberg J, Ooi HS, Nyegaard M. Identification of the BRD1 interaction network and its impact on mental disorder risk. Genome medicine. 2016 Dec;8(1):1–20.

47. Pralhada Rao R, Vaidyanathan N, Rengasamy M, Mammen Oommen A, Somaiya N, Jagannath MR. Sphingolipid metabolic pathway: an overview of major roles played in human diseases. Journal of lipids. 2013 Oct;2013.

48. Hernández-Corbacho MJ, Salama MF, Canals D, Senkal CE, Obeid LM. Sphingolipids in mitochondria. Biochimica et Biophysica Acta (BBA)-Molecular and Cell Biology of Lipids. 2017 Jan 1;1862(1):56–68.

49. Brignone MS, Lanciotti A, Camerini S, De Nuccio C, Petrucci TC, Visentin S, Ambrosini E. MLC1 protein: a likely link between leukodystrophies and brain channelopathies. Frontiers in cellular neuroscience. 2015 Apr 1;9:106.

50. Yang Y, Gocke AR, Lovett-Racke A, Drew PD, Racke MK. PPAR alpha regulation of the immune response and autoimmune encephalomyelitis. PPAR research. 2008;2008.

51. Walzer T, Chiossone L, Chaix J, Calver A, Carozzo C, Garrigue-Antar L, Jacques Y, Baratin M, Tomasello E, Vivier E. Natural killer cell trafficking in vivo requires a dedicated sphingosine 1-phosphate receptor. Nature immunology. 2007 Dec;8(12):1337–44.

52. Mayol K, Biajoux V, Marvel J, Balabanian K, Walzer T. Sequential desensitization of CXCR4 and S1P5 controls natural killer cell trafficking. Blood, The Journal of the American Society of Hematology. 2011 Nov 3;118(18):4863–71.

53. Walzer T, Chiossone L, Chaix J, Calver A, Carozzo C, Garrigue-Antar L, Jacques Y, Baratin M, Tomasello E, Vivier E. Natural killer cell trafficking in vivo requires a dedicated sphingosine 1-phosphate receptor. Nature immunology. 2007 Dec;8(12):1337–44.

54. Townsend MJ, Weinmann AS, Matsuda JL, Salomon R, Farnham PJ, Biron CA, Gapin L, Glimcher LH. T-bet regulates the terminal maturation and homeostasis of NK and Vα14i NKT cells. Immunity. 2004 Apr 1;20(4):477–94.

55. Orange JS. Natural killer cell deficiency. Journal of Allergy and Clinical Immunology. 2013 Sep 1;132(3):515–25.

56. Biron CA, Byron KS, Sullivan JL. Severe herpesvirus infections in an adolescent without natural killer cells. New England Journal of Medicine. 1989 Jun 29;320(26):1731–5.

57. Etzioni A, Eidenschenk C, Katz R, Beck R, Casanova JL, Pollack S. Fatal varicella associated with selective natural killer cell deficiency. The Journal of pediatrics. 2005 Mar 1;146(3):423–5.

58. Ornstein BW, Hill EB, Geurs TL, French AR. Natural killer cell functional defects in pediatric patients with severe and recurrent herpesvirus infections. The Journal of infectious diseases. 2013 Feb 1;207(3):458–68.

59. Ornstein BW, Hill EB, Geurs TL, French AR. Natural killer cell functional defects in pediatric patients with severe and recurrent herpesvirus infections. The Journal of infectious diseases. 2013 Feb 1;207(3):458–68.

60. Mace EM, Hsu AP, Monaco-Shawver L, Makedonas G, Rosen JB, Dropulic L, Cohen JI, Frenkel EP, Bagwell JC, Sullivan JL, Biron CA. Mutations in GATA2 cause human NK cell deficiency with specific loss of the CD56bright subset. Blood, The Journal of the American Society of Hematology. 2013 Apr 4;121(14):2669–77.

61. Meacci E, Garcia-Gil M. S1P/S1P receptor signaling in neuromuscolar disorders. International journal of molecular sciences. 2019 Jan;20(24):6364.

62. Kim TJ, Mitsutake S, Igarashi Y. The interaction between the pleckstrin homology domain of ceramide kinase and phosphatidylinositol 4, 5-bisphosphate regulates the plasma membrane targeting and ceramide 1-phosphate levels. Biochemical and biophysical research communications. 2006 Apr 7;342(2):611–7.

63. Kitatani K, Iwabuchi K, Snider A, Riboni L. Sphingolipids in inflammation: from bench to bedside. Mediators of inflammation. 2016 Jan 1;2016.

64. Karunakaran I, van Echten-Deckert G. Sphingosine 1-phosphate–A double edged sword in the brain. Biochimica et Biophysica Acta (BBA)-Biomembranes. 2017 Sep 1;1859(9):1573–82.

65. Drouillard A, Mathieu AL, Marçais A, Belot A, Viel S, Mingueneau M, Guckian K, Walzer T. S1PR5 is essential for human natural killer cell migration toward sphingosine-1 phosphate. Journal of Allergy and Clinical Immunology. 2018 Jun 1;141(6):2265–8.

66. Dickinson AJ, Meyer M, Pawlak EA, Gomez S, Jaspers I, Allbritton NL. Analysis of sphingosine kinase activity in single natural killer cells from peripheral blood. Integrative Biology. 2015 Apr 7;7(4):392–401.

67. Wang H, Liang S, Wang M, Gao J, Sun C, Wang J, Xia W, Wu S, Sumner SJ, Zhang F, Sun C. Potential serum biomarkers from a metabolomics study of autism. Journal of psychiatry & neuroscience: JPN. 2016 Jan;41(1):27.

68. Kurochkin I, Khrameeva E, Tkachev A, Stepanova V, Vanyushkina A, Stekolshchikova E, Li Q, Zubkov D, Shichkova P, Halene T, Willmitzer L. Metabolome signature of autism in the human prefrontal cortex. Communications biology. 2019 Jun 21;2(1):1–0.

69. Canals D, Perry DM, Jenkins RW, Hannun YA. Drug targeting of sphingolipid metabolism: sphingomyelinases and ceramidases. British journal of pharmacology. 2011 Jun;163(4):694–712.

70. Adam D, Wiegmann K, Adam-Klages S, Ruff A, Krönke M. A novel cytoplasmic domain of the p55 tumor necrosis factor receptor initiates the neutral sphingomyelinase pathway. Journal of Biological Chemistry. 1996 Jun 14;271(24):14617–22.

71. Kolevzon A, Bush L, Wang AT, Halpern D, Frank Y, Grodberg D, Rapaport R, Tavassoli T, Chaplin W, Soorya L, Buxbaum JD. A pilot controlled trial of insulin-like growth factor-1 in children with Phelan-McDermid syndrome. Molecular autism. 2014 Dec;5(1):1–9.

72. Fastman J, Foss-Feig J, Frank Y, Halpern D, Harony-Nicolas H, Layton C, Sandin S, Siper P, Tang L, Trelles P, Zweifach J. A Randomized Controlled Trial of Intranasal Oxytocin in Phelan-McDermid Syndrome.

73. Earls RH, Menees KB, Chung J, Gutekunst CA, Lee HJ, Hazim MG, Rada B, Wood LB, Lee JK. NK cells clear α-synuclein and the depletion of NK cells exacerbates synuclein pathology in a mouse model of α-synucleinopathy. Proceedings of the National Academy of Sciences. 2020 Jan 21;117(3):1762–71.

74. Togashi H, Sakisaka T, Takai Y. Cell adhesion molecules in the central nervous system. Cell adhesion & migration. 2009 Jan 1;3(1):29–35.

75. Muller D, Mendez P, DeRoo M, Klauser P, Steen S, Poglia L. Role of NCAM in spine dynamics and synaptogenesis. Structure and Function of the Neural Cell Adhesion Molecule NCAM. 2010:245–56.

