## Supplemental Figures for "Large deletions perturb peripheral transcriptomic and metabolomic profiles in Phelan-McDermid syndrome"

**Supplemental figures and legends**:

Supplemental Figure 1. Landscape of *SHANK3* sequence variants in the current study.

Supplemental Figure 2. Characterization of 22q13.3 breakpoints and disrupted genes.

Supplemental Figure 3. Overlap of differentially expressed genes at FDR < 5%.

Supplemental Figure 4. Direct protein-protein interaction (PPI) network.

Supplemental Figure 5. CD56+ NK cell enrichment gene set enrichment.

Supplemental Figure 6. CD56+ NK cell-specific expression via scRNA-seq.

Supplemental Figure 7. Gene expression on 22q13.3 that predicts *S1PR5* expression.

Supplemental Figure 8. Exploratory analysis of phenotype-transcriptome associations.

Supplemental Figure 9. Metabolites associated with Class II mutations.

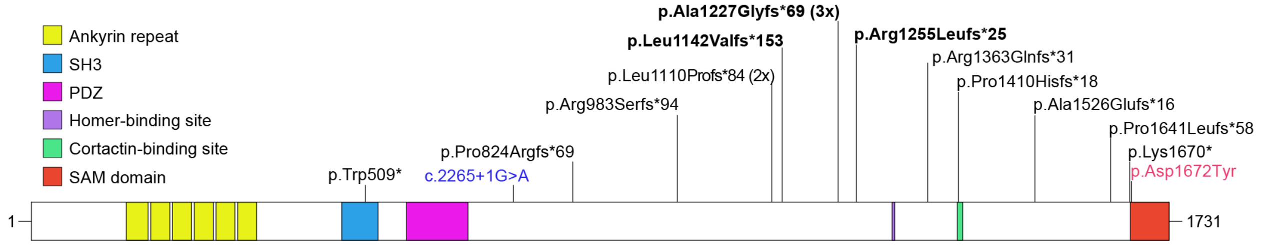


**Supplemental Figure 1. Landscape of *SHANK3* sequence variants in the current study.** Recurrent mutations are indicated in black, missense in red and splice site variants in blue. Protein domains are from UniProt; the homer and cortactin binding sites are indicated as previously reported.

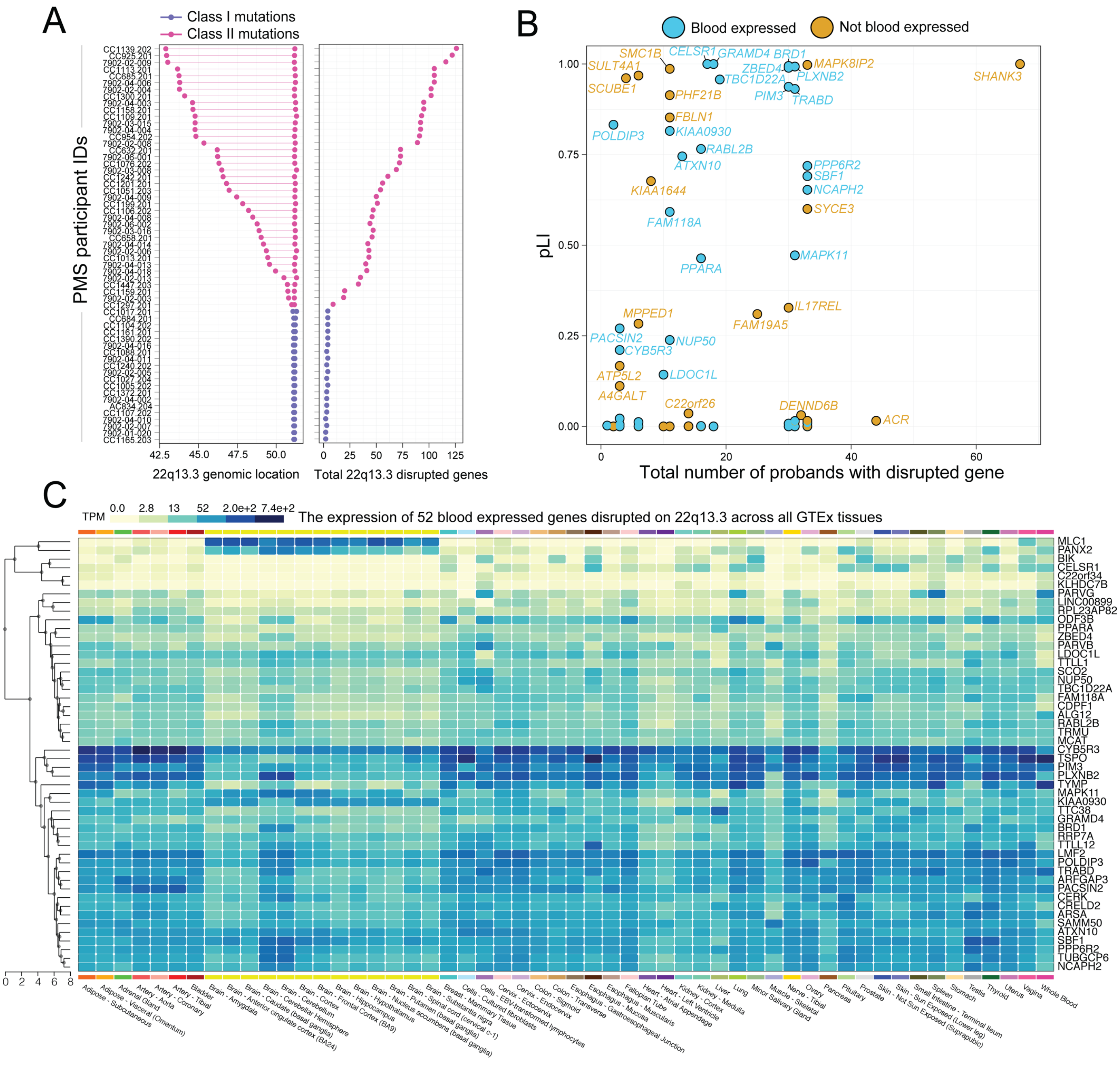


**Supplemental Figure 2. Characterization of 22q13.3 breakpoints and disrupted genes.** **(A)** 22q13.3 deletion sizes (left; x-axis) and the total number of disrupted genes (right; x-axis) for each PMS participant (y-axis) designated as Class I mutations (without sequence variants) and Class II mutations. (**B**) Probability of loss of function intolerance (pLI) scores computed for each of the disrupted genes (y-axis) relative to the frequency of a gene to be disrupted across all PMS participants with transcriptome data (x-axis, max value=68 probands). pLI scores close to 1 indicate gene intolerance to heterozygous and homozygous loss of function. A total of 16 blood expressed displayed pLI > 0.5 and 8 genes had a pLI > 0.9. (**C**) We identified 52 blood expressed genes spanning the single largest 22q13.3 deletion in the current study. The average expression values (TPM) of these 52 genes were plotted across 30 distinct tissues from the Geneotype-Tissue Expression (GTEx) consortium, and in some instances, covering multiple regions per tissue. White indicates low expression and dark blue indicates high expression. This plot was generated using the multi-gene query function in the GTEx browser.

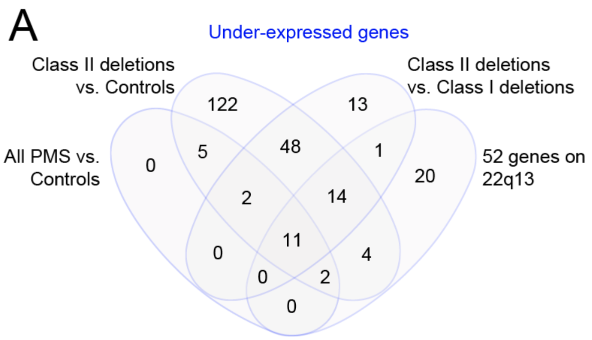


**Supplemental Figure 3. Overlap of differentially expressed genes at FDR < 5%**. The overlap of (**A**) under-expressed and (**B**) over-expressed genes for i) all PMS participants relative to controls, ii) Class II mutations relative to controls, and iii) Class II mutations relative to Class I mutations. We also examined the overlap of the 52 peripheral blood expressed genes on 22q13 encompassed within large Class II mutations in the current study.

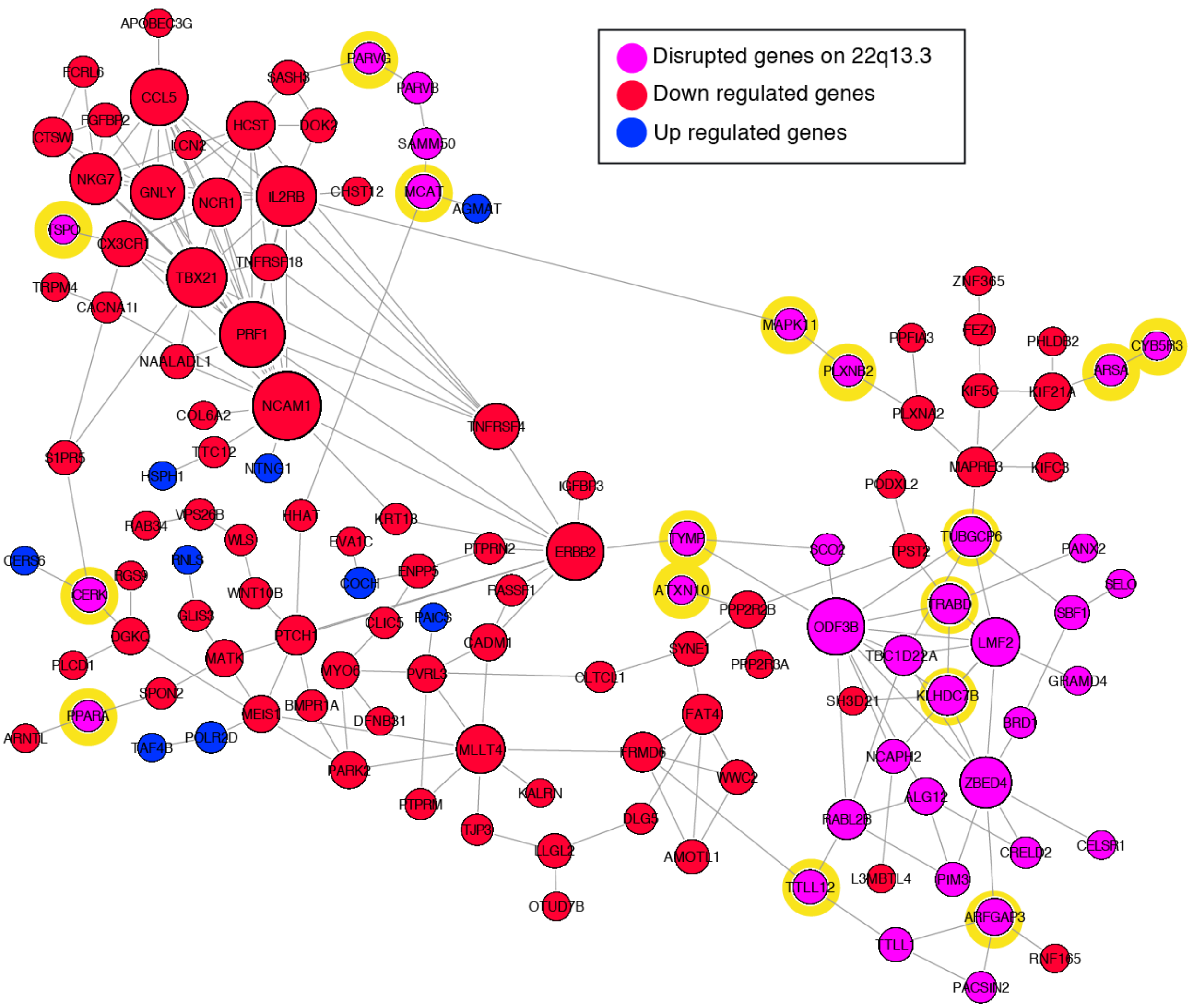


**Supplemental Figure 4. Direct protein-protein interaction (PPI) network.** All 52 genes on 22q13.3 and differentially expressed genes (FDR < 5%) associated with PMS participants with Class II mutations were tested for enrichment of direct PPIs. The network contained significantly higher connectivity than expected by chance (*p* <1.0e-16). Nodes are colored by under-expressed genes (red), over-expressed genes (blue), and disrupted genes on 22q13.3 (pink). Yellow background is given to genes on 22q13.3 found to interact with differentially expressed genes.

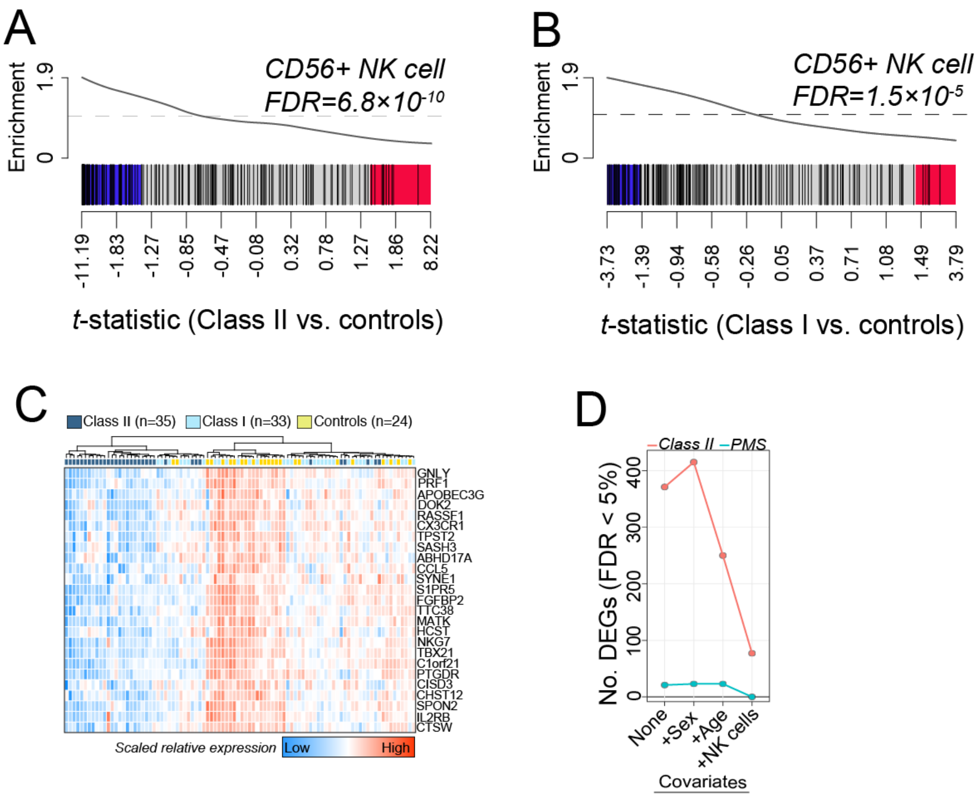


**Supplemental Figure 5. CD56+ NK cell enrichment gene set enrichment**. CAMERA gene-set enrichment results for differentially expressed genes associated with (**A**) Class II mutations and (**B**) Class I mutations. Enrichment was tested for 190 genes that are differentially expressed CD56+ NK cells compared to all other cell types in the scRNA-seq experiment. (**C**) Unsupervised clustering of 25 CD56+ NK cell-specific genes distinguishes 82% (n=29) of Class II mutations from remaining samples. (**D**) The total number of significant differentially expressed genes in participants with Class II mutations (FDR < 5%) after adjusting for different covariates, reveals adjusting for CD56+ NK cell frequencies results in loss of ~69% of Class II-related DEGs.

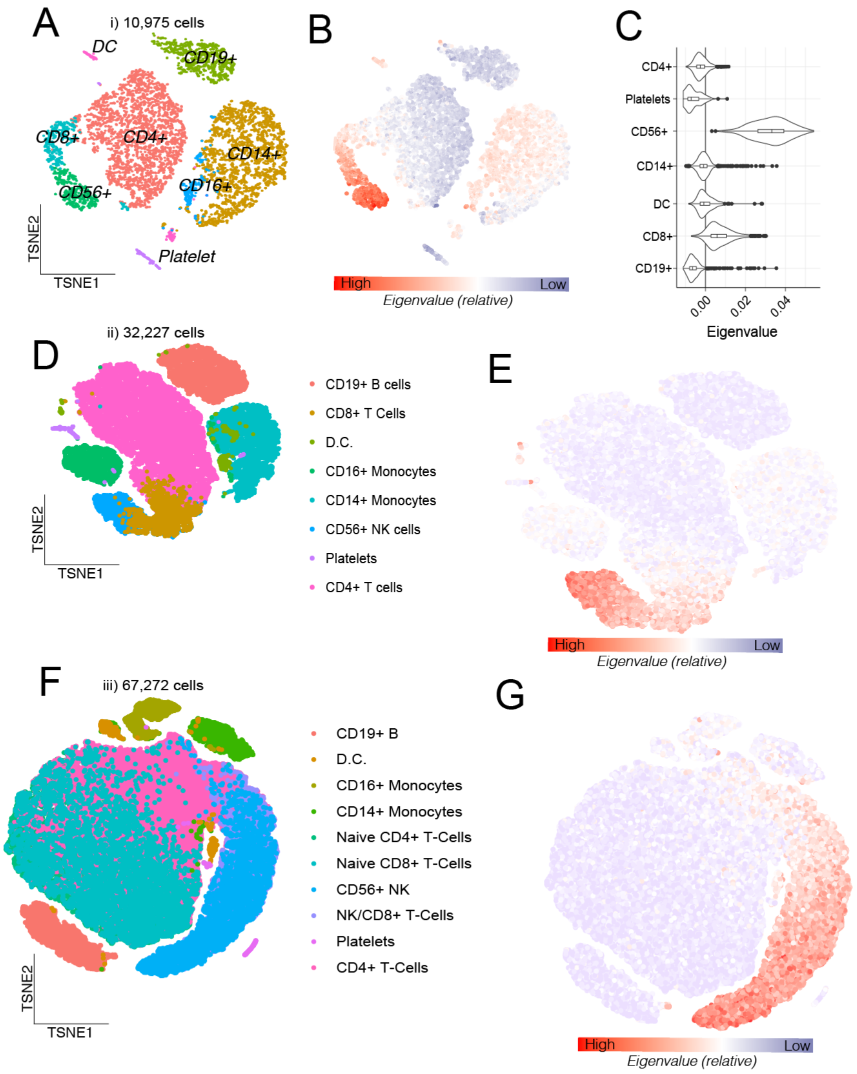


**Supplemental Figure 6. CD56+ NK cell-specific expression via scRNA-seq**. TSNE clustering and cell type identification of eight main immune cell types across three independent studies: (**A**) the first dataset comprised of 10,975 PBMCs (v2 Chemistry); (**D**) the second dataset comprised of 33,227 PBMCs (v2 Chemistry), both were downloaded from the list of publically available 10X Genomic Inc. datasets; (**F**) third data set was comprised of 67,272 PBMCs and was obtained from Zheng et al., 2017[28]. Next, the normalized and scaled scRNA-seq expression data was used to create an eigenvalue (per cell) of 208 significantly under-expressed genes in participants with Class II mutations, which was projected onto each TSNE and color coded to illustrate high expression of these genes in CD56+ NK cells (blue=low, red=high) (**B, E, G**, respectively).(**C**) For clarity, eigenvalues (x-axis) were plotted as a violin plot for each cell type (y-axis) to illustrate strength of enrichment (merging CD14+ and CD16+ monocytes).

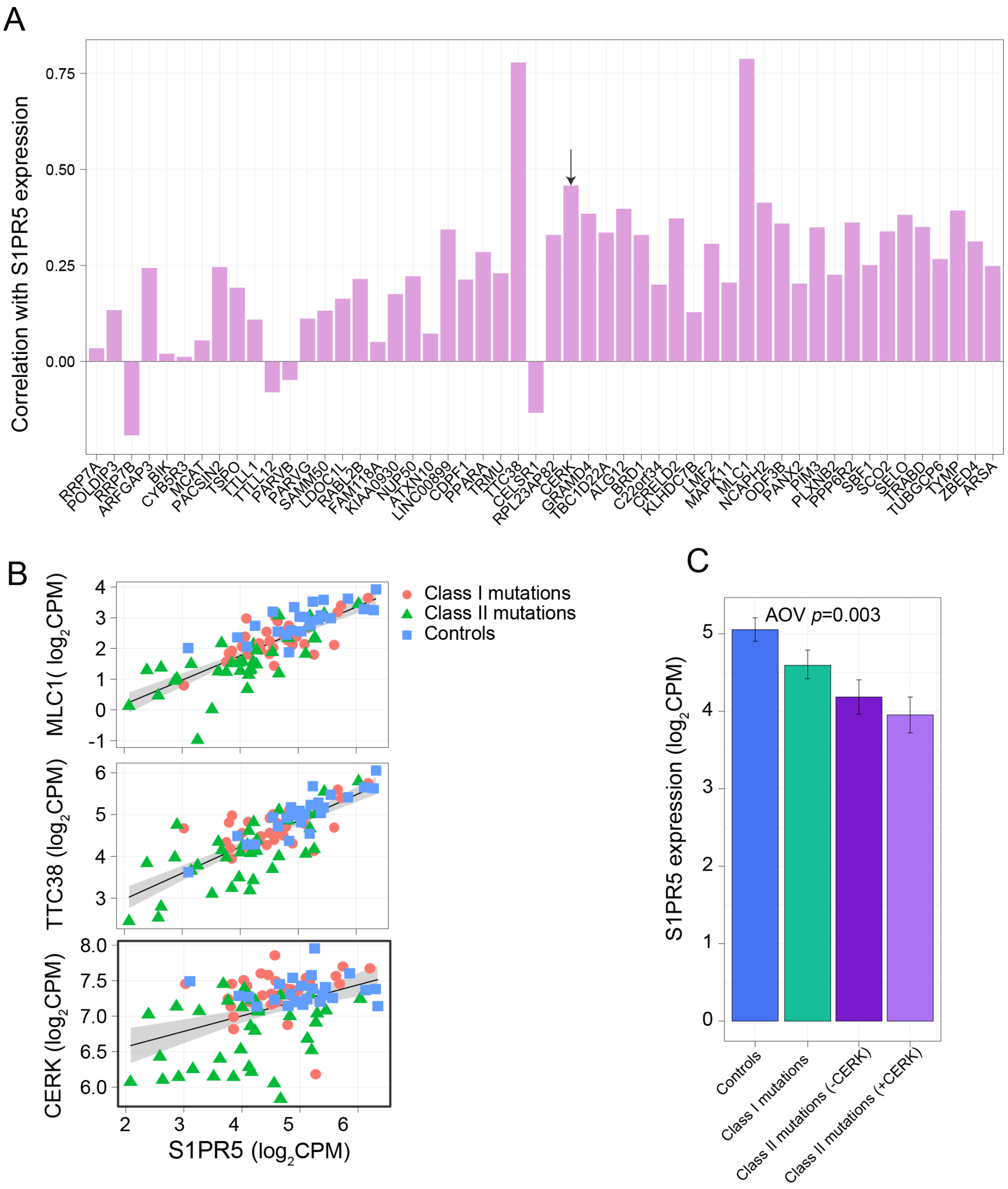


**Supplemental Figure 7. Gene expression on 22q13.3 that predicts *S1PR5* expression**. (**A**) Barplots depicting the Pearson's correlation coefficient (y-axis) between gene expression of the 52 blood expressed genes on 22q13.3 number relative to *S1PR5* expression. *CERK* is denoted with an arrow. (**B**) The top three genes on 22q13.3 with the highest associations (y-axis) with *S1PR5* expression (x-axis) are depicted. (**C**) We anticipated that by parsing PMS participants with Class II mutations spanning *MLC1*, *TTC38*, and *CERK*, respectively, that those individuals would display lower expression of *S1PR5* relative to the remaining of individuals with Class II mutations. We found that only participants with Class II mutations spanning *CERK* were predictive of *S1PR5* expression, in that reduced expression of this gene was evident when compared with the remaining Class II mutations. An analysis of variance (AOV) was used to test for significance.

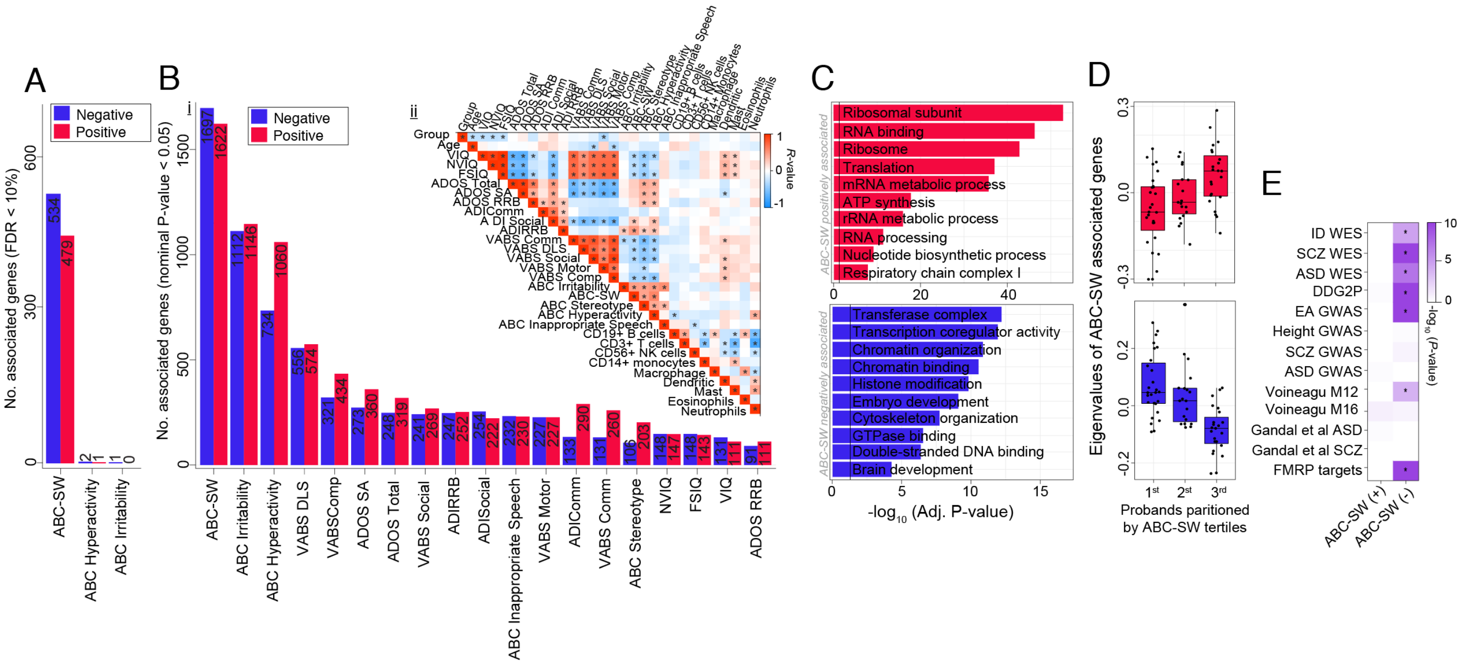


**Supplemental Figure 8. Exploratory analysis of phenotype-transcriptome associations**. Barplots depicting the total number of genes positively (red) and negatively (blue) associated with each clinical measure presented in **Table 1** according to (**A**) a FDR < 10% and (**Bi**) a nominal *p*-value < 0.05. (**Bii**) Pearson's correlation matrix among all clinical traits in the current study (red=high; blue=low; *=significant association). (**C**) Functional annotation of genes positively and negatively associated with ABC-lethargy (social withdrawal). (**D**) To conceptualize these associations, all positively and negatively associated genes were summarized into one singular value using singular value decomposition, respectively. Probands were partitioned into tertiles according to ABC-lethargy scores and the resulting eigenvalues were plotted across low ($1^{st\ tertile}$) to high ($3^{rd\ tertile}$) scores confirming significant positive and negative associations. (**E**) Gene set enrichment analysis shows a significant enrichment of disease risk genes for intellectual disability (ID), schizophrenia (SCZ), autism spectrum disorder (ASD) and educational attainment (EA) among genes negatively associated with ABC-lethargy. Significance was calculated using a Fisher's exact test relative to a genome background of genes expressed in the current study.

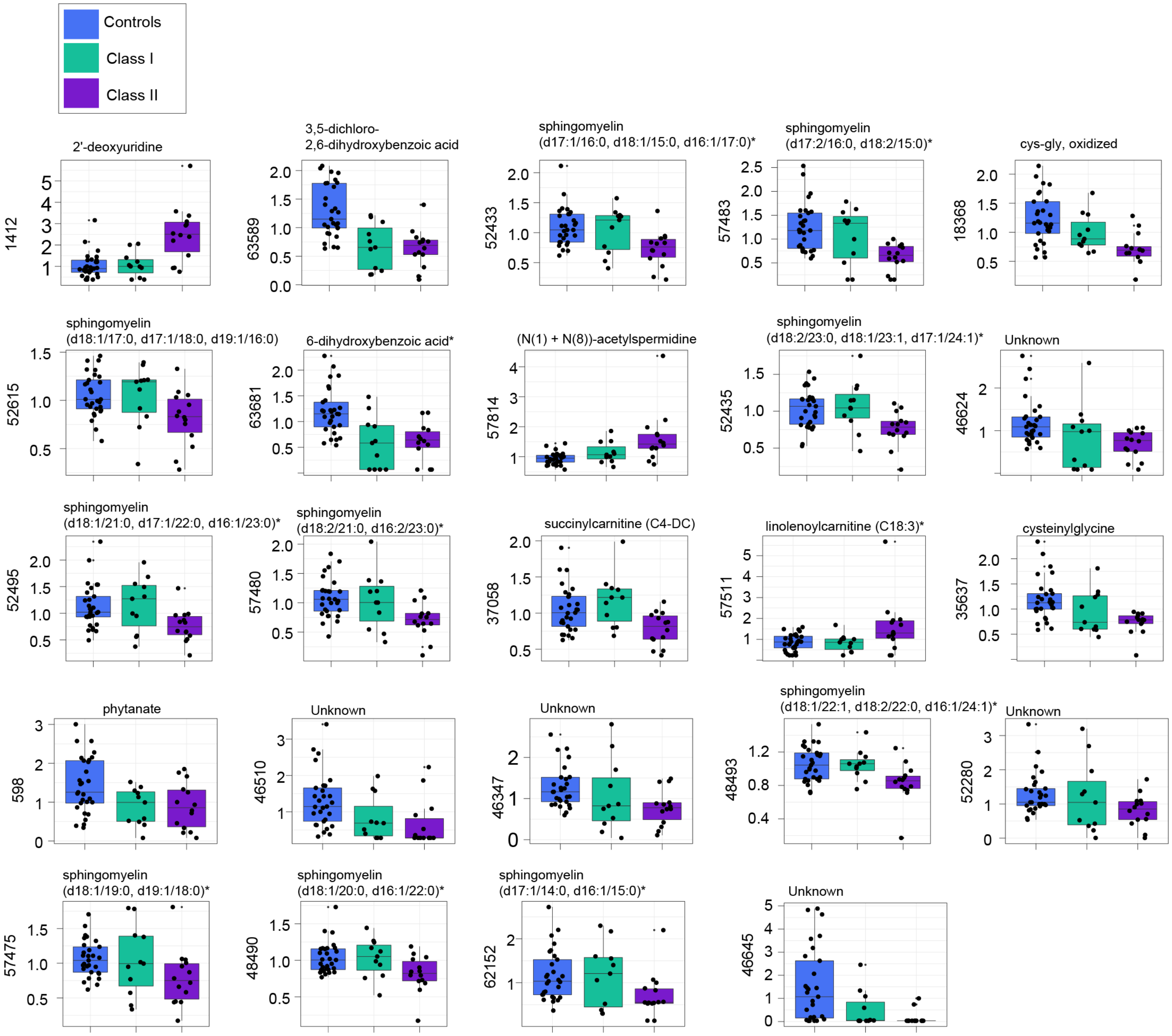


**Supplemental Figure 9. Metabolites associated with Class II mutations**. Twenty-four differentially abundant metabolites significantly associated with Class II mutations relative to controls (FDR < 10%) are displayed. Scaled metabolite abundance (y-axes) was partitioned by deletion group (x-axes). The y-axis labels indicate compound identifiers and the main titles indicate the biochemical identifiers.
